## Supporting Information for "A network modelling approach to assess non-pharmaceutical disease controls in a worker population: An application to SARS-CoV-2"

**3 Supporting Text S3: Parameterisation of contact risk** **12**

**4 Supporting Text S4: Non-intervention scenario calibration** **13**

**5 Additional figures** **15**

**6 Additional tables** **26**

### 1 Supporting Text S1: Network generation

#### 1.1 Workplace contacts

We describe here the procedure for generating the static workplace contacts in our network model. The static workplace contacts are generated using a 'configuration model' style algorithm [1], allowing the specification of a desired degree distribution based on pre-pandemic conditions (i.e. 100% attendance at work). This differs to an Erdös-Rényi random graph generation [2], which assumes a Poisson degree distribution. We adapted the standard configuration model to allow a variable amount of clustering, where a higher value of clustering led to more contacts being made within a workplace compared to between different workplaces. We set the probability of making contact with an individual in another workplace compared to an individual within the same workplace at 0.05. Unlike the standard configuration model, we did not allow edges to be made with oneself or repeated edges.

Iterating over each workplace, the steps defining our algorithm were:

1. Draw a random degree for each node in the workplace from the appropriate distribution - these form a number of 'half-edges' for each node.
2. Limit the number of half-edges per node to be no more than the size of the workplace.
3. While there are at least two unconnected half-edges:
   (a) Pick an unconnected half-edge at random;
   (b) With sector-specific probability, connect chosen half-edge to a random node outside its own workplace, but within the same sector, forming an edge;
   (c) Otherwise: pick another unconnected half-edge at random from the workplace and connect the two, forming an edge (with the condition that this does not create an edge with oneself or a repeated edge);
   (d) If the above conditions can not be satisfied, the chosen half-edge is discarded from the network.
4. If a single unconnected half-edge remains, it is discarded from the network.
5. To incorporate those now working from home, we only include an edge between two nodes in the final network if both nodes have returned to work.

#### 1.2 Social contacts

We describe here the procedure for generating social contacts in our network model, first involving the grouping of individuals into friendship cliques, followed by sampling each timestep the contacts made amongst those in the friendship group.

##### Establishing friendship groups

We created friendship groups for each individual using a configuration model [1] to allow the specification of a desired degree distribution. As in the workplace contact layer, we adapted the standard configuration model to allow for greater clustering (with a probability of 0.5 that each contact was made with a friend of a friend rather than someone at random) and did not allow edges with oneself or repeated edges.

Iterating over each individual, the steps defining our algorithm were:

1. Draw a random degree for all nodes in the network - these form half-edges for each node and represent the number of friends each person has.
2. Limit the number of half-edges per node to a specified maximum, set to 100.
3. While there are at least two unconnected half-edges:
   (a) Pick an unconnected half-edge at random;
   (b) Find friends-of-friends for chosen node, limited to those with unconnected half-edges that are not already friends with chosen node, or oneself;
   (c) If the number of friends-of-friends satisfying the above conditions is nonzero, pick a node at random from friends-of-friends and form an edge;
   (d) Otherwise: pick a half-edge at random from the whole population and connect to form an edge

Note, in step 3(b) someone who was listed as being friends with more than one friend of the target node was included in the friends-of-friend group multiple times. Thus, in step 3(c), that individual was more likely to be chosen to form a connection with.

##### Generation of daily social contacts

We generated the daily dynamic social contacts for each timestep of the simulation. While there were still nodes unassigned to a cluster of social contacts on the given timestep, we applied the following steps:

1. Choose an unassigned node at random.
2. Generate a cluster size from the appropriate workday/non-workday distribution.
3. Find the number of unassigned friends of the chosen node and limit the cluster size to this number.
4. Assign friends to the cluster at random and create contacts with the chosen node.
5. For each node in the cluster:
   (a) Make contacts with the other nodes if they are friends;
   (b) Otherwise, replace the contact with a friend from outside the cluster.
6. Repeat steps 1-5 until all nodes are assigned.

### 2 Supporting Text S2: Network parameterisation

#### 2.1 Workplace contacts

We used the Warwick Social Contact Survey [3–5] to parameterise the degree distributions for both static and dynamic contacts occurring in workplaces. We considered all contacts reported to occur at 'Work/School', limited to those in the following occupations:

- Entertainment;
- Health;
- Labour;
- Mechanic;
- Office;
- Public;
- Research;
- Service;
- Teaching;
- Transport.

We further split the 'Service' occupation into the following specific occupations:

- Retail
- Post office
- Accommodation
- Hospitality
- Bank
- Real estate
- Vet
- Travel agent
- Cleaner
- Sports
- Hairdresser
- Funeral director

We limited static contacts to those that took place 4+ days a week (highest frequency in survey). We limited dynamic contacts to those records specifying that the contact had occurred 'for the first time today' (lowest frequency in survey). We did not include contacts with frequency between these extremes.

Across occupations, the distributions for the number of work related contacts were generally heavy tailed. We found lognormal distributions had a reasonable correspondence with the empirical data for both static contacts (Figs. S1 and S2) and dynamic contacts (Figs. S3 and S4). Among the disaggregated ‘Service’ occupations, some occupations had very little data and we were unable to obtain a fit. In these instances, we instead used parameters fit to the aggregated ‘Service’ occupation (Table S1).

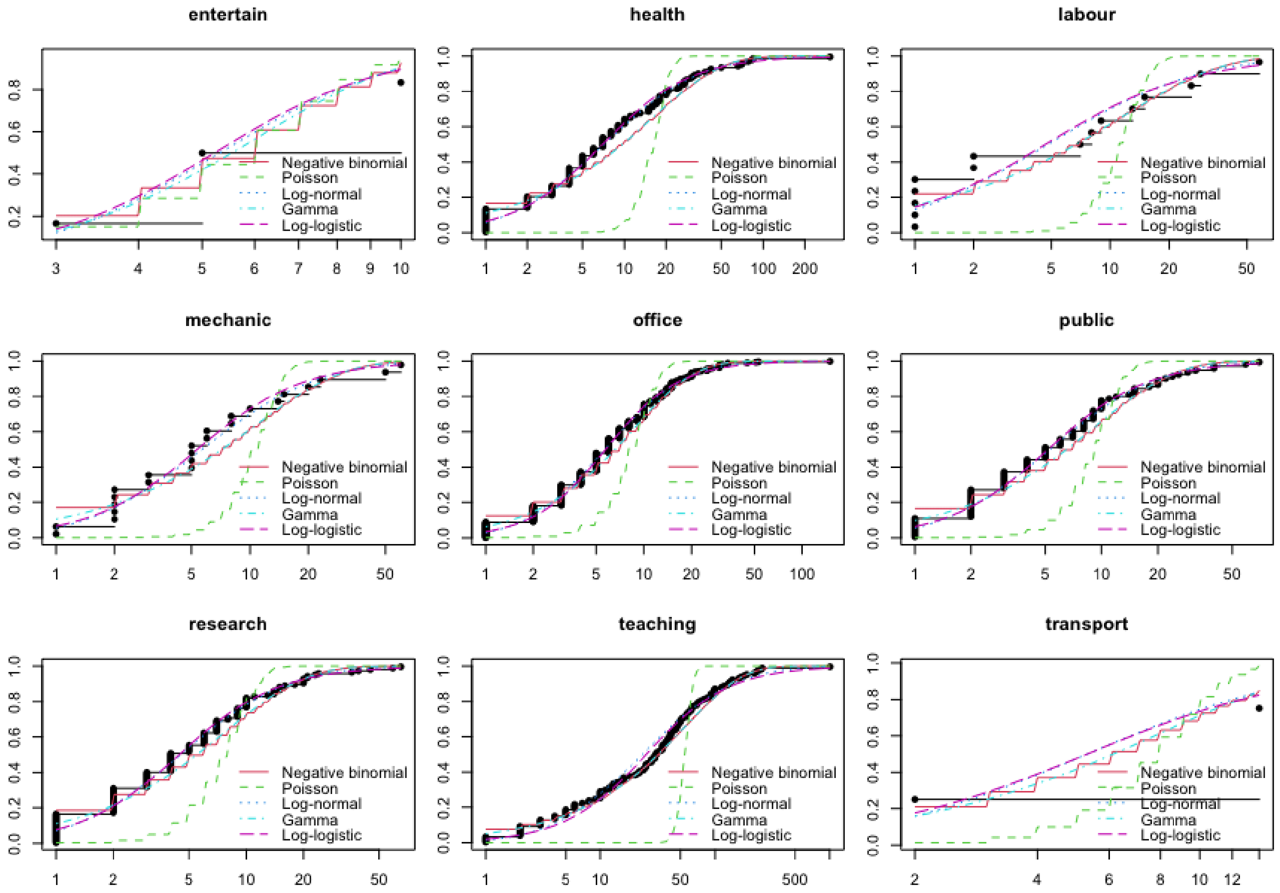

**Fig. S1: Cumulative distribution functions from parametric fits to the number of typical (static) work associated contacts for non-service occupations.** We present the empirical cumulative distribution function (black line and dots), estimated from the Social Contact Survey. We also display cumulative distribution functions from four parametric distributions fit to the data: negative binomial (solid red line); Poisson (green dashed line); lognormal (blue dotted line); gamma (green dot-dash line); log-logistic (purple dashed line).

In the final step, we mapped each of the 41 work sectors obtained from the ONS data to an occupation provided in the Social Contact Survey (Table S2). In other words, for the network parameterisation of workplace contacts in a given ONS sector, we used the fit distribution for the linked occupation from the Social Contact Survey.

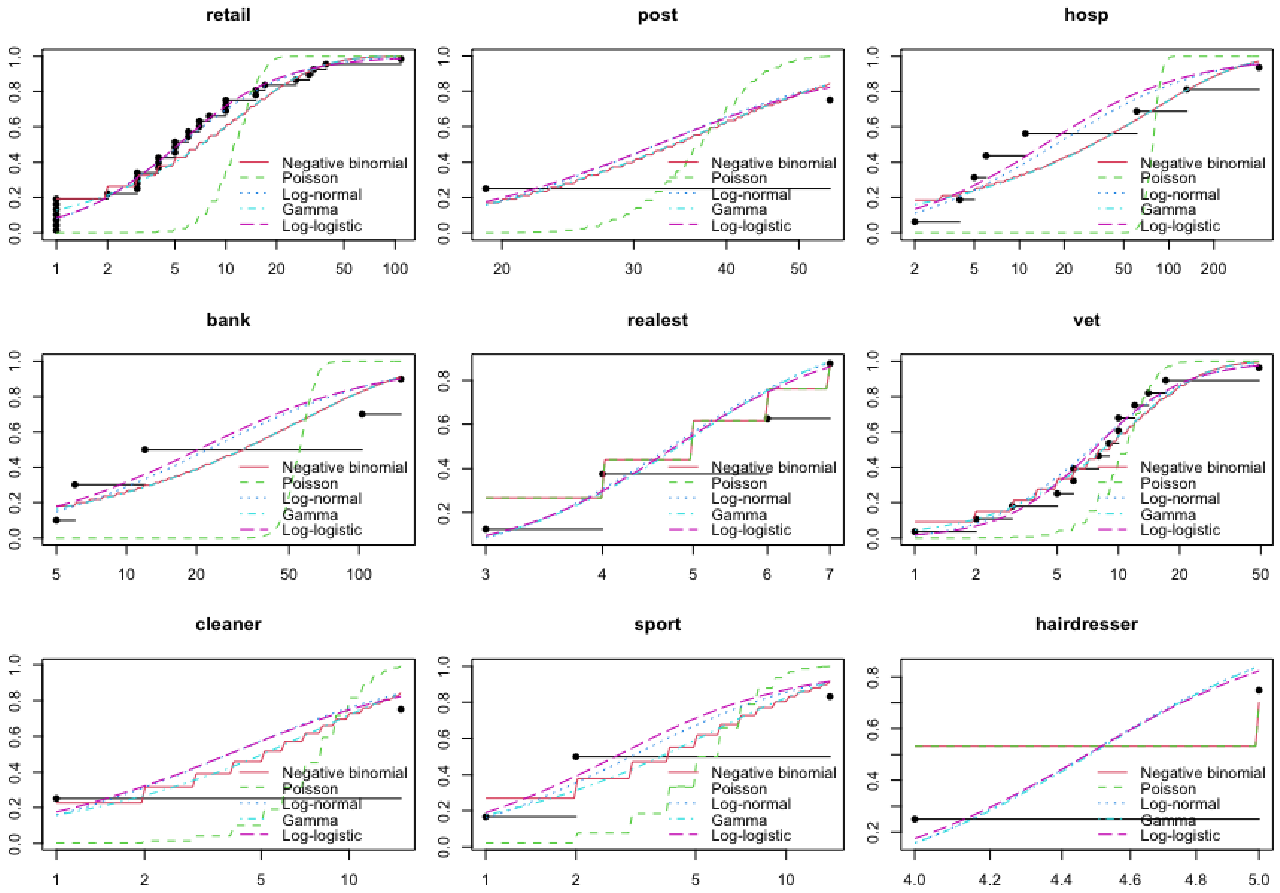

**Fig. S2: Cumulative distribution functions from parametric fits to the number of typical (static) work associated contacts for service based occupations.** We present the empirical cumulative distribution function (black line and dots), estimated from the Social Contact Survey. We also display cumulative distribution functions from four parametric distributions fit to the data: negative binomial (solid red line); Poisson (green dashed line); lognormal (blue dotted line); gamma (green dot-dash line); log-logistic (purple dashed line).

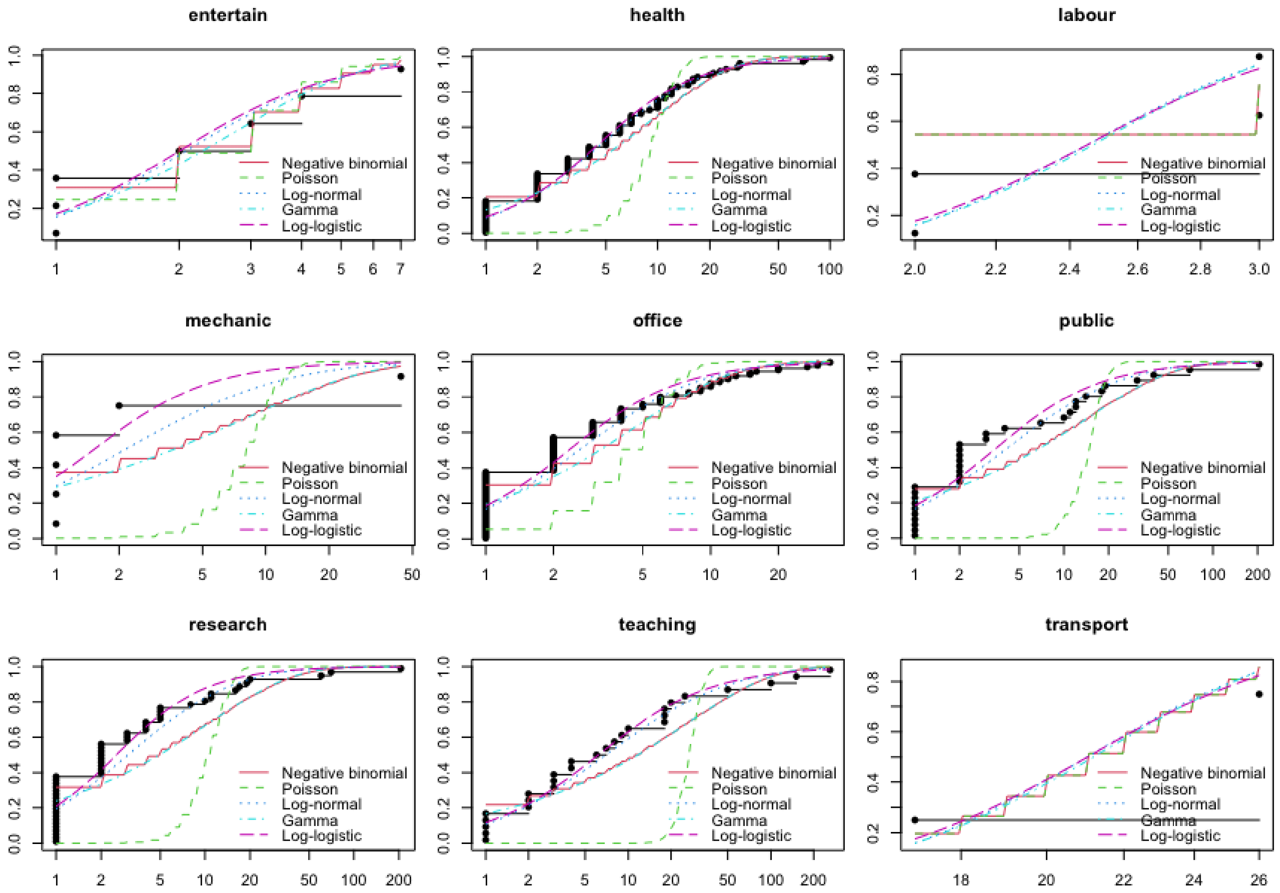

**Fig. S3: Cumulative distribution functions from parametric fits to the number of dynamic work associated contacts for non-service occupations.** We present the empirical cumulative distribution function (black line and dots), estimated from the Social Contact Survey. We also display cumulative distribution functions from four parametric distributions fit to the data: negative binomial (solid red line); Poisson (green dashed line); lognormal (blue dotted line); gamma (green dot-dash line); log-logistic (purple dashed line).

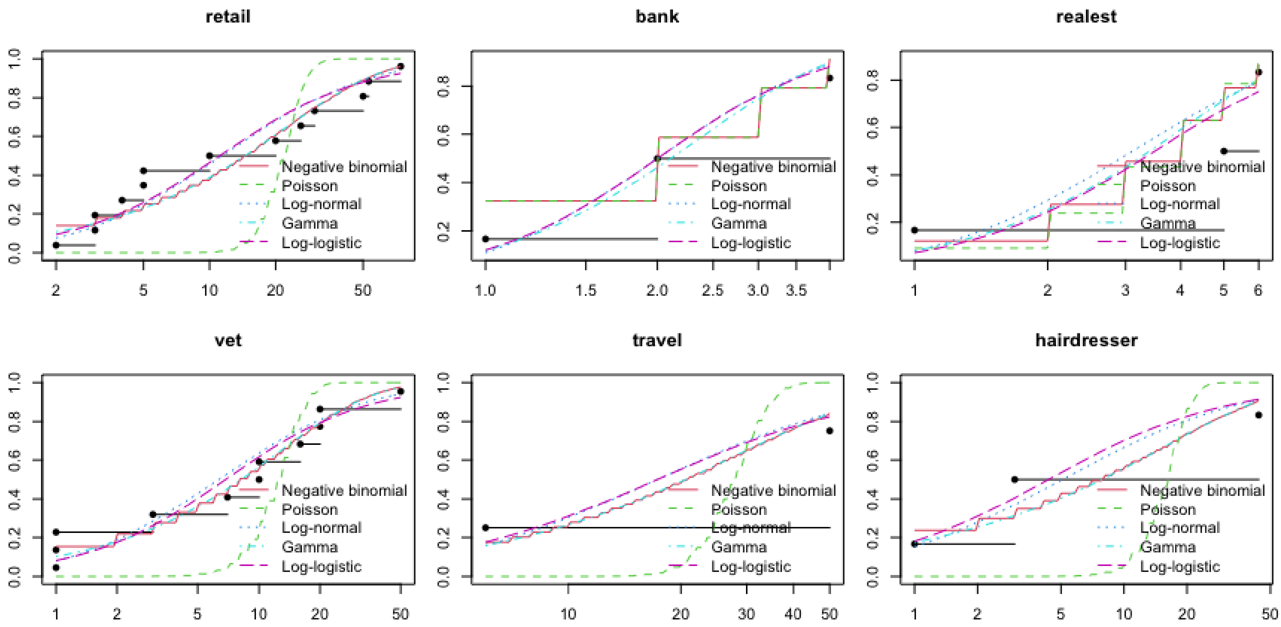

**Fig. S4: Cumulative distribution functions from parametric fits to the number of dynamic work associated contacts for service based occupations.** We present the empirical cumulative distribution function (black line and dots), estimated from the Social Contact Survey. We also display cumulative distribution functions from four parametric distributions fit to the data: negative binomial (solid red line); Poisson (green dashed line); lognormal (blue dotted line); gamma (green dot-dash line); log-logistic (purple dashed line).

**Table S1:** Description of the lognormal distributions acquired by maximum likelihood estimation by fitting to the number of work related contacts in the specified occupations, estimated from Social Contact Survey records. All values are given to 2d.p. For service based occupations, entries with value $---$ had an insufficient number of records to allow a parametric fit to the data to be performed. In these instances, we used the inferred distributions for the overall service occupation.

| | **Static** | | **Dynamic** | |
|---|---|---|---|---|
| **Occupation** | **Meanlog** | **SDlog** | **Meanlog** | **SDlog** |
| Entertainment | 1.67 | 0.49 | 0.73 | 0.72 |
| Health | 1.96 | 1.25 | 1.54 | 1.12 |
| Labour | 1.57 | 1.40 | 0.90 | 0.20 |
| Mechanic | 1.73 | 1.09 | 0.75 | 1.40 |
| Office | 1.73 | 0.91 | 0.96 | 0.99 |
| Public | 1.67 | 1.04 | 1.39 | 1.43 |
| Research | 1.50 | 1.02 | 1.11 | 1.29 |
| Teaching | 3.15 | 1.43 | 1.94 | 1.56 |
| Transport | 1.67 | 0.97 | 3.05 | 0.21 |
| Overall non-service | 1.94 | 1.22 | 1.26 | 1.22 |
| Retail | 1.69 | 1.20 | 2.40 | 1.20 |
| Post office | 3.48 | 0.53 | — | — |
| Accommodation | — | — | — | — |
| Hospitality | 2.86 | 1.78 | — | — |
| Bank | 3.10 | 1.44 | 0.69 | 0.57 |
| Real estate | 1.56 | 0.33 | 1.13 | 0.81 |
| Vet | 1.98 | 0.92 | 1.85 | 1.31 |
| Travel agent | — | — | 2.85 | 1.06 |
| Cleaner | 1.35 | 1.35 | — | — |
| Sports | 1.11 | 1.12 | — | — |
| Hairdresser | 1.50 | 0.11 | 1.63 | 1.59 |
| Funeral director | — | — | — | — |
| Overall service | 1.97 | 1.35 | 1.76 | 1.42 |
| Overall | 1.94 | 1.23 | 1.33 | 1.26 |

**Table S2:** The mappings applied for each of the 41 ONS sectors to a Social Contact Survey occupation.

| ONS working sectors | Social contact survey occupation group |
|---|---|
| Agriculture | Labour |
| Mining | Labour |
| Manufacturing (food) | Labour |
| Manufacturing (other) | Labour |
| Utilities and Waste | Labour |
| Construction | Labour |
| Motor Trade | Service - Retail |
| Wholesale | Service - Retail |
| Retail | Service - Retail |
| Transport | Transport |
| Transport Support | Transport |
| Postal | Service - Post |
| Accommodation | Service - Accommodation |
| Restaurant/Bar | Service - Hospitality |
| Broadcasting and Communication | Office |
| IT | Office |
| News | Office |
| Banking/Accounting | Service - Bank |
| Real Estate | Service - Real estate |
| Professional/Sci/Tech | Research |
| Veterinary | Service - Vet |
| Rental Companies | Service - Real estate |
| Employment/HR | Office |
| Travel Agency | Service - Travel agent |
| Security | Public |
| Cleaning | Service - Cleaner |
| Office (other) | Office |
| Public/Admin/Defence | Public |
| Education | Teaching |
| Hospital/Doctor/Dental | Health |
| Care Homes | Health |
| Social Work | Health |
| Arts | Entertainment |
| Betting | Entertainment |
| Sports | Service - Sports |
| Theme Parks | Entertainment |
| Religious Organisations | Public |
| Repair | Mechanic |
| Hairdressers | Service - Hairdresser |
| Funerals | Service - Funeral |
| Personal Services | Office |

#### 2.2 Friendship group size

We defined the distribution of friendship group sizes by scaling up the contacts recorded in the Warwick Social Contact Survey. We assumed that each response represented a typical day and the individual would contact everyone in the friendship group each week. Thus, for each individual $i$, we estimated the total number of friends they had, $N^i$, by calculating the sum

$$N^i_{\text{friends}} = n^i_{\text{daily}} + 3n^i_{\text{2-3 per week}} + 7n^i_{\text{once per week}} + 7n^i_{\text{less than once per week}},$$

where $n^i_{\text{daily}}$ represented the number of contacts individual $i$ had with friends that occurred every day, $n^i_{\text{2-3 per week}}$ the number of contacts with friends that occurred two to three times per week, $n^i_{\text{once per week}}$ the number of contacts with friends occurring only once per week, and $n^i_{\text{less than once per week}}$ the number of contacts with friends occurring at a frequency of less than once a week.

#### 2.3 Social workday and non-workday contacts

To parameterise workday and non-workday social contacts, we defined workdays to be days on which a count above zero of 'Work / School' contacts had been recorded. Otherwise, we treated it as a non-work day. For contacts in the social setting, we limited records to those specifying contacts that occurred regularly (not for the first time), were recorded as 'Leisure', 'Shopping' or 'Other' and either lasted longer than ten minutes or involved touching the other person. Similarly to the number of daily contacts in the workplace setting, the number of daily social contacts displayed a heavy tail. Once more, lognormal distributions agreed favourably with the empirical data (Fig. S5).

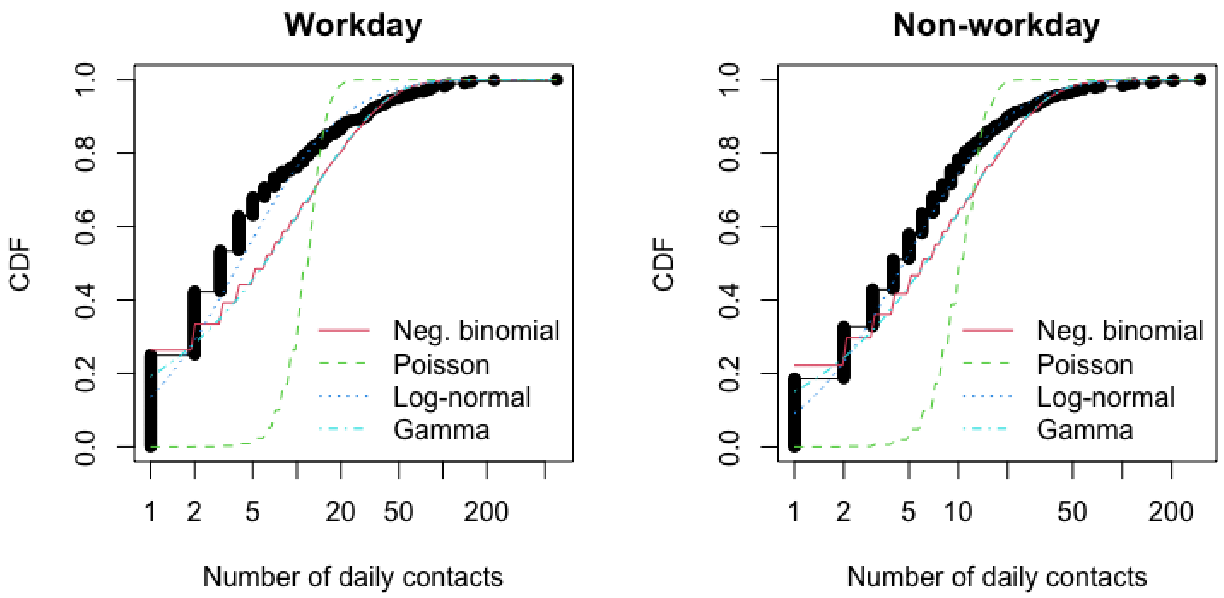

**Fig. S5: Parametric fits to the number of daily social contacts on workdays and non-workdays. (Left)** Workday social contacts; **(Right)** non-workday social contacts. In each panel, we present the empirical cumulative distribution function (black line and dots), estimated from the Social Contact Survey. We also display cumulative distribution functions from four parametric distributions fit to the data: negative binomial (solid red line); Poisson (green dashed line); lognormal (blue dotted line); gamma (green dot-dash line). The lognormal distribution had the strongest resemblance with the empirical data.

#### 3 Supporting Text S3: Parameterisation of contact risk

Each data record contained in the Warwick Social Contact Survey [3–5] included a field on the the duration of the interaction and whether it involved physical touch. We used these data attributes to scale the transmission risk of contacts occurring in non-household settings relative to household contacts. Explicitly, the contact survey found 80% of household contacts to involve touch. As a result, touch contacts contributed 80% to the household secondary attack rate estimate. The remaining 20% of the household secondary attack rate we attributed to non-touch contacts with a duration classification above 0; of the non-touch contacts, 80% had duration classified above 0.

We computed the relative transmission risk across static and dynamic work contacts for each occupation in the Social Contact Survey that had been used to parameterise the contact networks Table S1. We scaled transmission risks relative to the central estimate of overall unadjusted secondary attack rate in the household setting, with value 0.37 [6] (Table S3). The relative magnitude of those estimates, versus the household setting, were then used to scale the associated standard deviations. Using our mapping of occupations from the Social Contact Survey to each of the 41 work sectors (Table S2), for a given work sector we applied the transmission risk estimates associated with the linked occupation.

For transmission risks across social contacts, the above procedure returned a value of 0.2627. The relative magnitude of that estimate was used to scale the standard deviation, consequently set at 0.0213. For each individual, we drew the transmission potential across contacts in each respective setting from normal distributions with the specified mean and standard deviation values.

**Table S3:** Relative transmission risks across static and dynamic work contacts for those occupations in the Social Contact Survey we had used to parameterise the contact networks. We scaled transmission risks relative to the central estimate of overall unadjusted secondary attack rate in the household setting, with value 0.37 [6]. All values are stated to 2d.p.

| Occupation | Static transmission risk | Dynamic transmission risk |
|---|---|---|
| Entertainment | 0.13 | 0.27 |
| Health | 0.21 | 0.29 |
| Labour | 0.10 | 0.12 |
| Mechanic | 0.11 | 0.36 |
| Office | 0.13 | 0.18 |
| Public | 0.14 | 0.26 |
| Research | 0.11 | 0.15 |
| Teaching | 0.23 | 0.16 |
| Transport | 0.22 | 0.24 |
| Retail | 0.16 | 0.04 |
| Post | 0.13 | 0.18 |
| Accommodation | 0.20 | 0.18 |
| Hospitality | 0.41 | 0.18 |
| Bank | 0.44 | 0.10 |
| Real estate | 0.10 | 0.16 |
| Vet | 0.13 | 0.10 |
| Travel | 0.10 | 0.03 |
| Cleaner | 0.09 | 0.18 |
| Sport | 0.04 | 0.18 |
| Hairdresser | 0.16 | 0.41 |
| Funeral | 0.10 | 0.37 |

### 4 Supporting Text S4: Non-intervention scenario calibration

#### Model parameterisation

In the absence of isolation and contact tracing, we calibrated the system to have a 7-day moving average $R_t$ of approximately three in the early phase of the outbreak. We achieved this magnitude of spread by applying a scaling factor of 0.8 to the baseline transmission risk across a contact in each setting. For each replicate, we drew the probability of a case being asymptomatic from a Uniform(0.5,0.8) distribution and the relative infectiousness of an asymptomatic from a Uniform(0.3,0.7) distribution.

All simulations were run on a network size of 10,000 nodes. We ran batches of 100 stochastic simulations on different network configurations, as well as a run of 1,000 simulations whose realisations consisted of 50 separate networks (with 20 runs performed using each network). All individuals began susceptible, with the exception of ten individuals seeded in an infectious state; we set between five to eight individuals as being asymptomatically infected (randomly sampled), with the remaining individuals (between two to five) symptomatic.

#### Results summary

Across our simulated collection of mean generation times the median value was roughly seven days (Fig. S6). In the absence of interventions, a mean generation time estimate in the region of seven days corresponds with findings from an analysis of transmission pair data from mainland China by Ali *et al.* [7], which found serial intervals were on average 7.8 days in mid-January 2020 (prior to the implementation of nonpharmaceutical interventions).

For the early stages of the outbreak, we obtained a median 7-day moving average $R_t$ of about 3 (with the 50% prediction interval in the range 2 to 4), with considerable variation between runs. Approximately 60-90% of the overall population became infected over the course of the outbreak. The peak of the outbreak occurred after approximately two months. The outbreak duration was variable, usually concluding after 3-5 months. Workplace contacts (combining static and dynamic types) contributed the most to infection, followed by the social setting, households and lastly other random contacts. Temporally, infection spread and peaked at similar times across settings (Fig. S7).

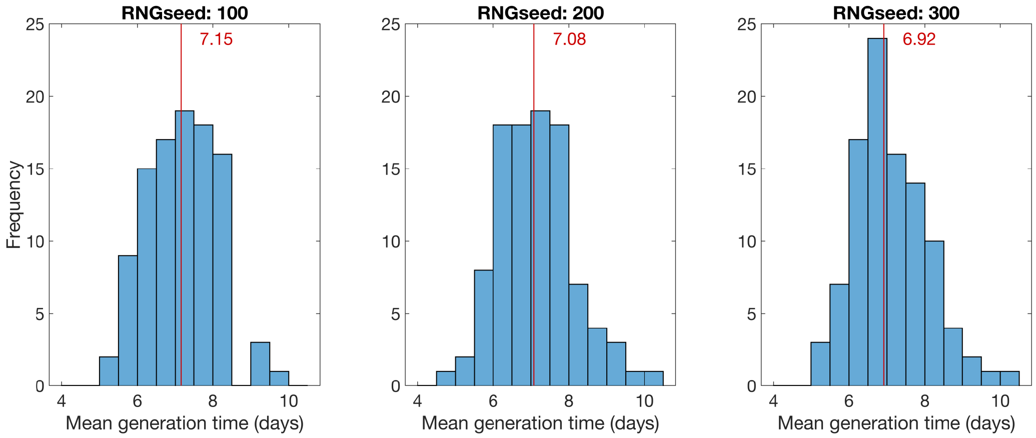

**Fig. S6: Distribution of mean generation times from 100 replicates.** Each panel was produced with the random number generator passed a different seed. The vertical line designates the median value, with the corresponding value (to 2 d.p.) stated alongside the line. We obtained a median value for the mean generation time of roughly seven days.

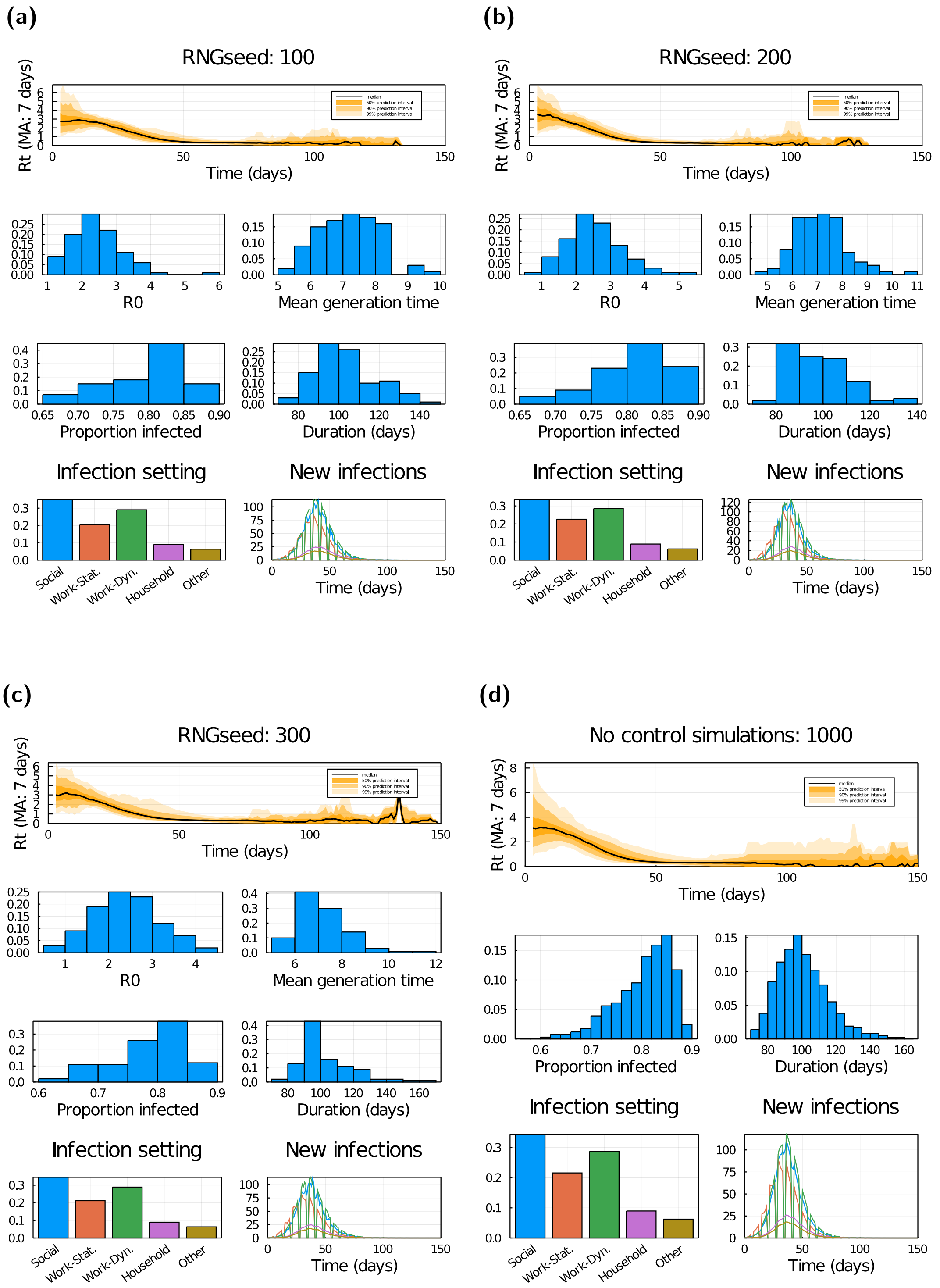

**Fig. S7: Outbreak temporal profiles and associated probability distributions in the absence of interventions. (a-c)** Estimated from three separate batches of 100 stochastic simulations, each on a different network realisation. *(Row one)* Daily $R_t$ estimate using a 7-day moving average. *(Row two, left)* Average number of individuals infected by the initial ten nodes. *(Row two, right)* Mean generation time. *(Row three, left)* Proportion of individuals infected over the course of the outbreak. *(Row three, right)* Duration of the outbreak. *(Row four, left)* Proportion of infections occurring in each setting. *(Row four, right)* Average (mean) amount of new daily infections in each setting. **(d)** Outputs summarised from 1,000 simulations (consisting of 50 network realisations and 20 runs per network). Panels match those presented in rows one, three and four for **(a-c)**.

#### 5 Additional figures

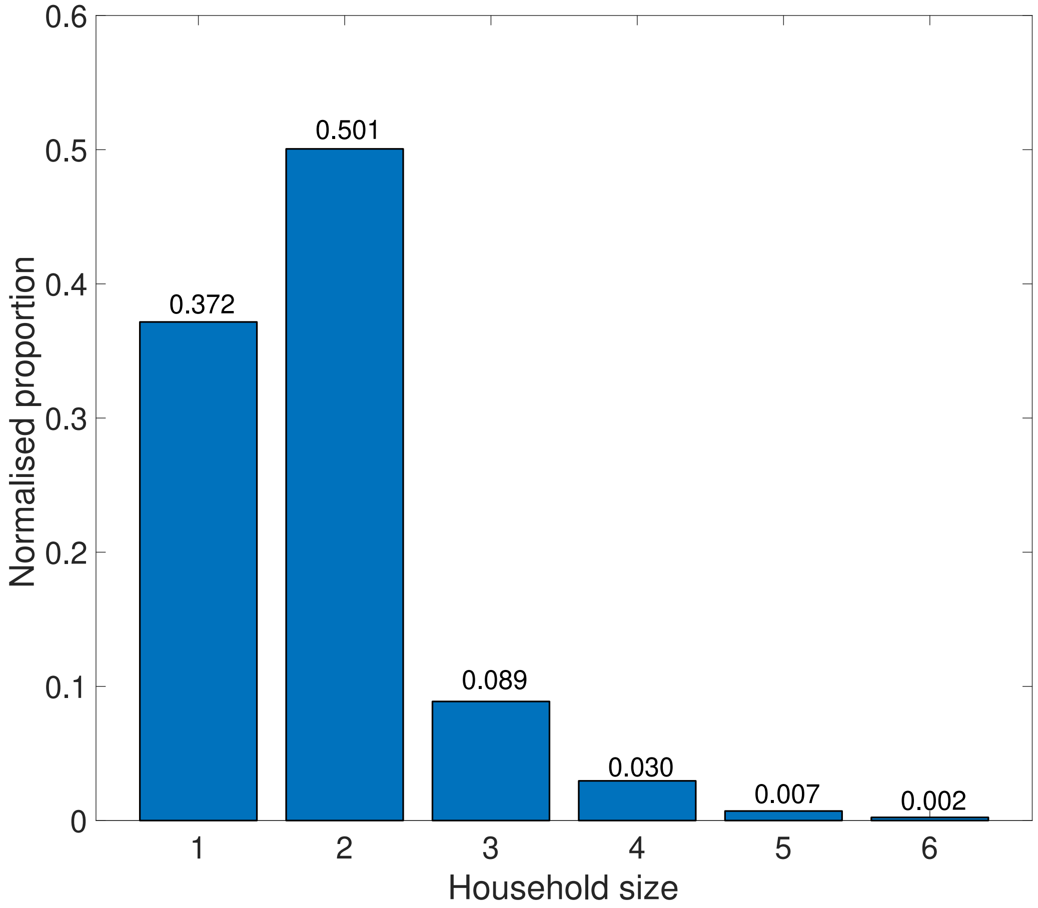

**Fig. S8: Normalised UK household distribution.** We calculated the proportion of households containing 1 to 6+ people between the ages of 20 - 70. When sampling from this distribution, we restricted the maximum household size to six people in an attempt to reduce the amount of overestimation of the number of active workers mixing within households, which results from the assumption that everyone in a household is an active worker. Normalised proportions for each household size are stated above the associated bar.

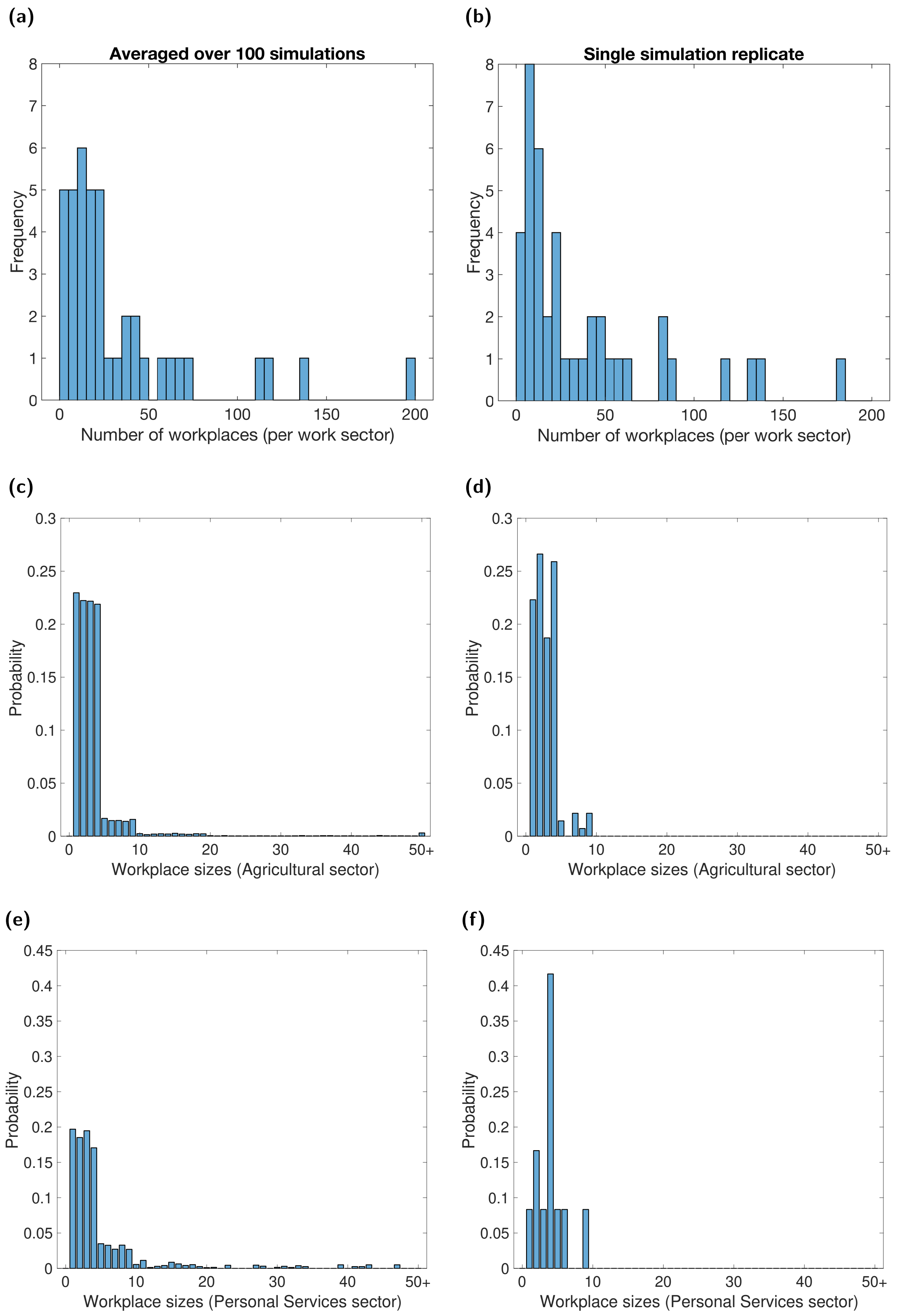

**Fig. S9: Number of workplaces and workplace sizes per sector produced by the network generation process.** In panels **(a,b)**, we show distributions of the amount of workplaces assigned to a work sector: **(a)** averaged across 100 simulations replicates; **(b)** for a single realisation. In panels **(c,d)**, we show probability mass functions for the size of workplaces generated for the 'Agriculture' sector: **(c)** averaged across 100 simulations replicates; **(d)** for a single realisation. We display equivalent information in panels **(e,f)** for the 'Personal Services' sector.

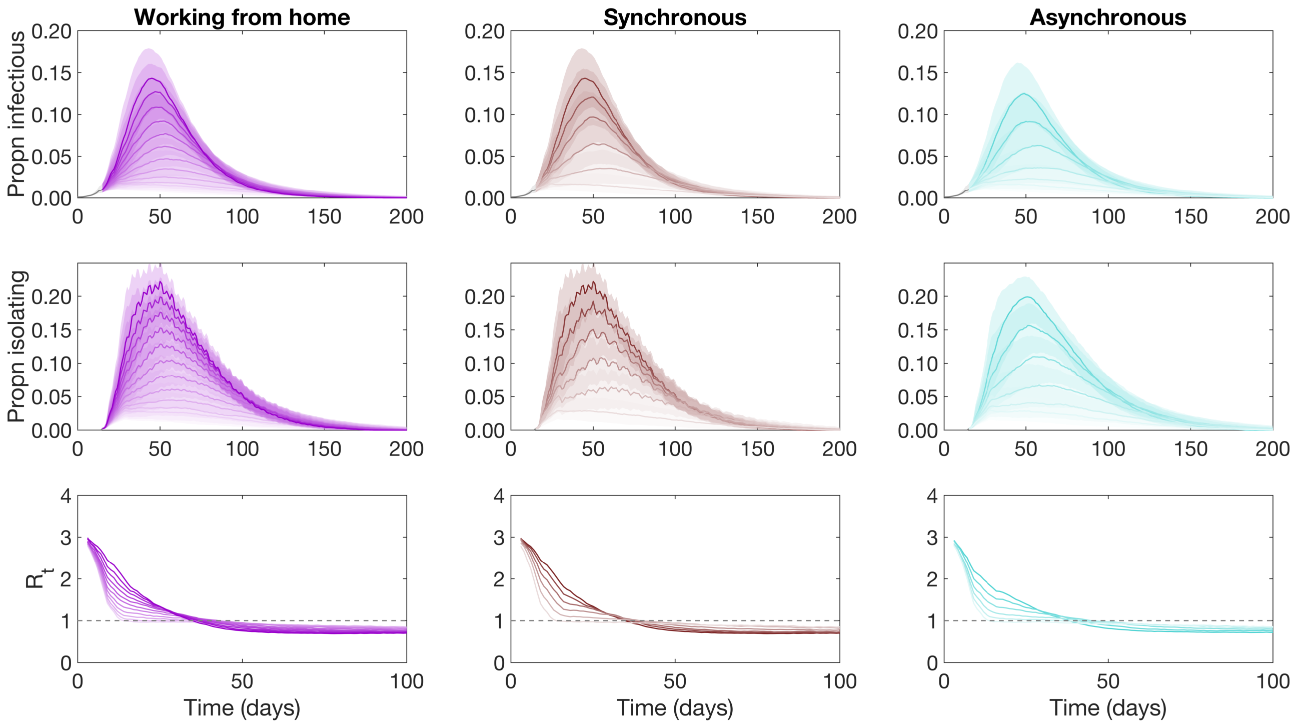

**Fig. S10: Infectious case prevalence, isolation and $R_t$ temporal profiles under alternative worker practices and scheduling.** We considered three work practice and scheduling assumptions: **(left column)** proportion of workers that work from home; **(central column)** synchronous work pattern; **(right column)** asynchronous work pattern. From day 15, test, trace and isolate guidance was introduced, with an adherence of 70%. For the statistics **(row one)** infectious case prevalence, **(row two)** proportion in isolation, and **(row three)** the effective reproduction number $R_t$ (the number of people, on average, each person that became infected at time $t$ passed the virus onto), we present median temporal traces (solid lines), with the shaded regions in rows one and two representing the 50% prediction intervals. Lighter intensities correspond to: a higher fraction of workers working from home (ranging from 0 to 1; left column); a greater number of days per week being spent working from home rather than spent at the workplace (ranging from 0 days to 5 days; central and right column).

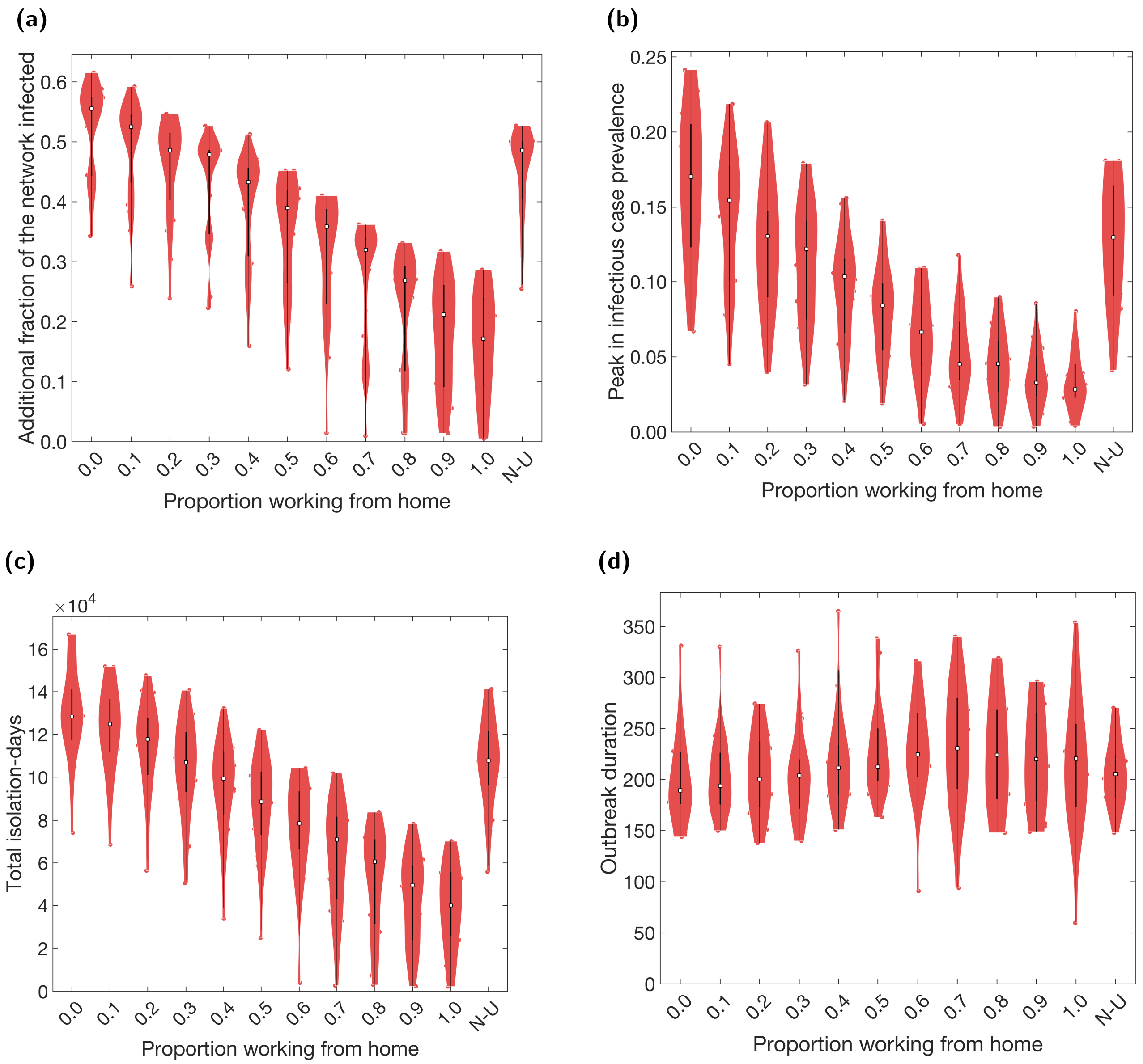

**Fig. S11: Case and isolation summary statistics under differing fractions of workers working from home using a single network structure.** We introduced NPIs from day 15 onwards, with varying proportions of the workforce working from home. N-U corresponds to non-uniform proportions working from home across the work sectors (see Table 5). We summarise outputs from 20 simulations on a single network realisation. We assumed an adherence of 70% in all runs. The white markers denote medians and solid black lines span the 25th to 75th percentiles. **(a)** Additional proportion of the population that were infectious post introduction of NPIs (day 15 onwards). **(b)** Peak in infectious case prevalence. **(c)** Total isolation-days. **(d)** Outbreak duration. We observe a large amount of variability in outcomes, which results from differences in epidemiological properties between simulations runs, such as the distribution of initial infections, the asymptomatic probability, and the relative infectiousness of an asymptomatic case (all of which were randomly generated at the start of each simulation).

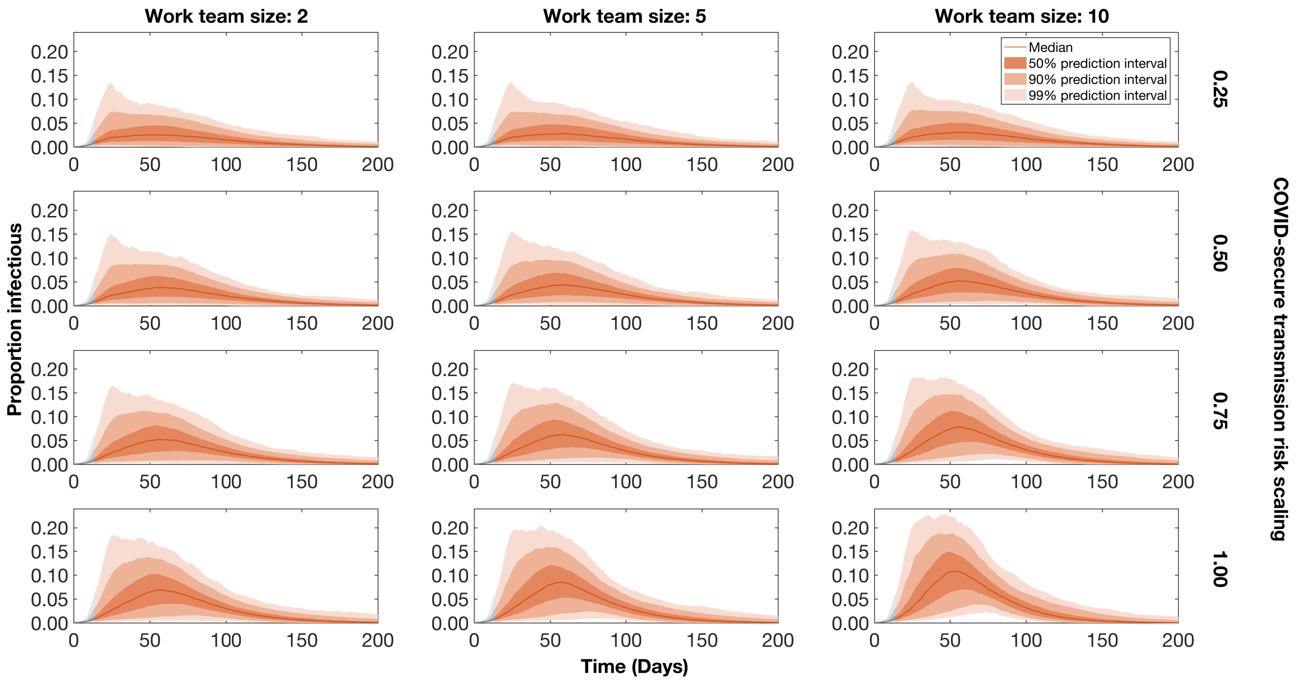

**Fig. S12: Temporal profiles of the proportion of the population in an infectious state for twelve combinations of COVID-secure workplace parameters.** We display outputs for combinations of, from day 15, work team sizes being capped at 2, 5 or 10 people, paired with scaling the transmission risk in COVID-secure workplaces by 0.25, 0.50, 0.75 or 1.00, respectively. Also from day 15, trace, trace and isolate measures were introduced and had an adherence percentage of 70%. We assumed all individuals were at the workplace five days a week (Monday to Friday). Traces and regions in grey correspond to the period where no interventions were in place (up to day 15). The solid line gives the median trace. Filled regions depict the 50%, 90% and 99% prediction intervals (with dark, moderate and light shading, respectively).

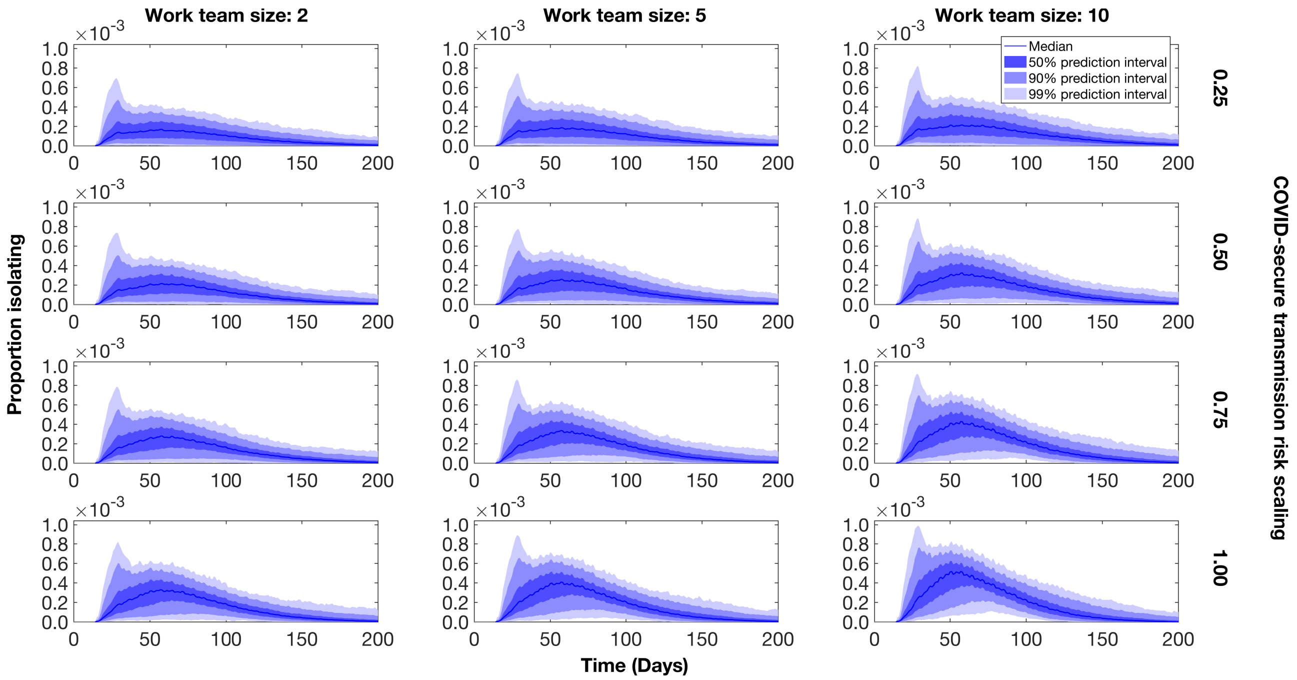

**Fig. S13: Temporal profiles of the proportion of the population in isolation under twelve combinations of COVID-secure workplace parameters.** We display outputs for combinations of, from day 15, work team sizes being capped at 2, 5 or 10 people, paired with scaling the transmission risk in COVID-secure workplaces by 0.25, 0.50, 0.75 or 1.00, respectively. Also from day 15, trace, trace and isolate measures were introduced and had an adherence percentage of 70%. We assumed all individuals were at the workplace five days a week (Monday to Friday). Traces and regions in grey correspond to the period where no interventions were in place (up to day 15). The solid line gives the median trace. Filled regions depict the 50%, 90% and 99% prediction intervals (with dark, moderate and light shading, respectively).

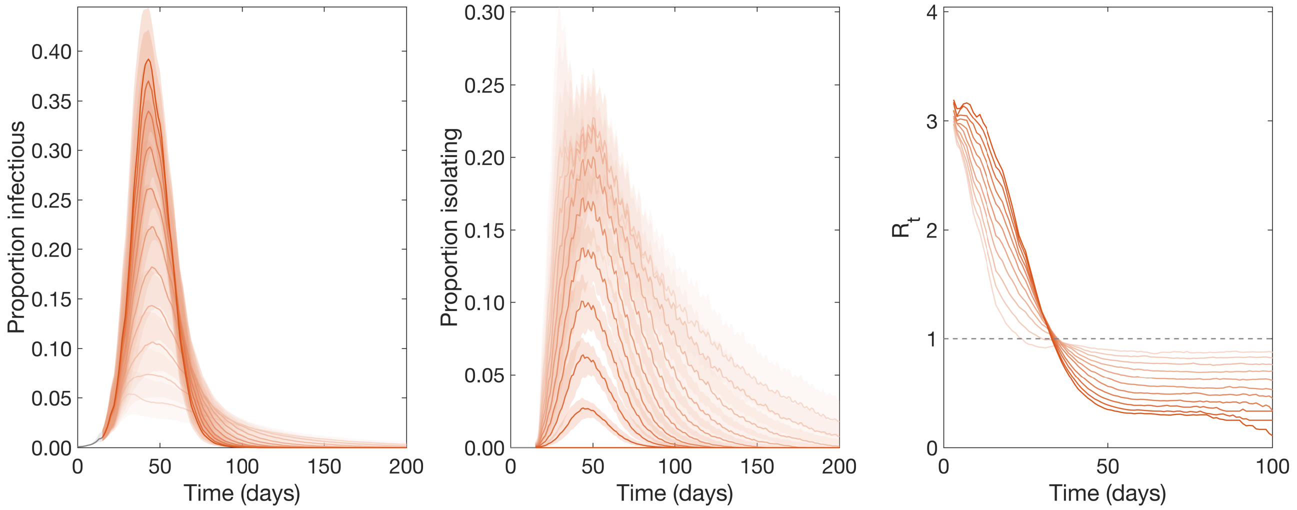

**Fig. S14: Infectious case prevalence, isolation and $R_t$ temporal profiles under self-isolation and contact tracing NPIs.** For the statistics **(column one)** infectious case prevalence, **(column two)** proportion in isolation, and **(column three)** the effective reproduction number $R_t$ (the number of people, on average, each person that became infected at time $t$ passed the virus onto), we present median temporal traces (solid lines), with lighter intensity corresponding to a higher adherence to the interventions (ranging from 0 to 1). In columns one and two, shaded regions represent 50% prediction intervals.

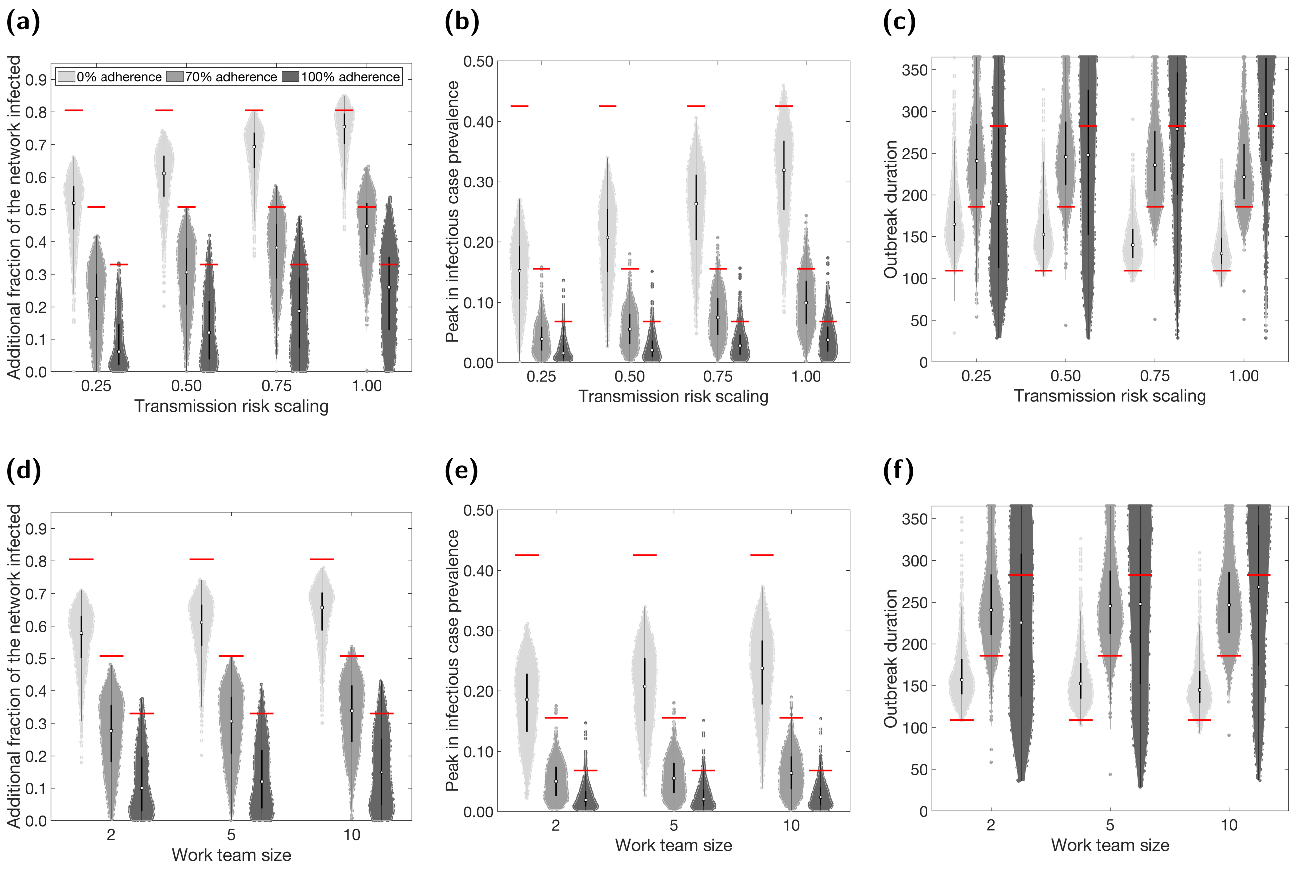

**Fig. S15: COVID-secure workplace measures and sensitivity to test, trace and isolation adherence of public health measurable quantities.** We introduced NPIs from day 15 onwards, alongside COVID-secure workplace measures. We compare three scenarios of adherence to isolation and test-and-trace measures: 0% (lightest shaded violins); 70% adherence (moderate shaded violins); 100% (darkest shaded violins). In panels **(a-c)** we fixed the work team size at 5 and varied the relative scaling of transmission risk under COVID-secure conditions. In panels **(d-f)** we fixed the relative scaling of transmission risk at 0.5 and varied the work team size. We summarise outputs from 1,000 simulations (with 20 runs per network, for 50 network realisations). The white markers denote medians and solid black lines span the 25th to 75th percentiles. The horizontal red lines represent the median value attained for the given adherence value to test, trance and isolation in the absence of any other COVID-secure workplace measures. We present the following summary statistics: **(a,d)** Additional proportion of the population that were infectious post introduction of NPIs (day 15 onwards). **(b,e)** Peak in infectious case prevalence. **(c,f)** Outbreak duration. We give central and 95% prediction intervals for each summary statistic distribution in Tables S7&S8.

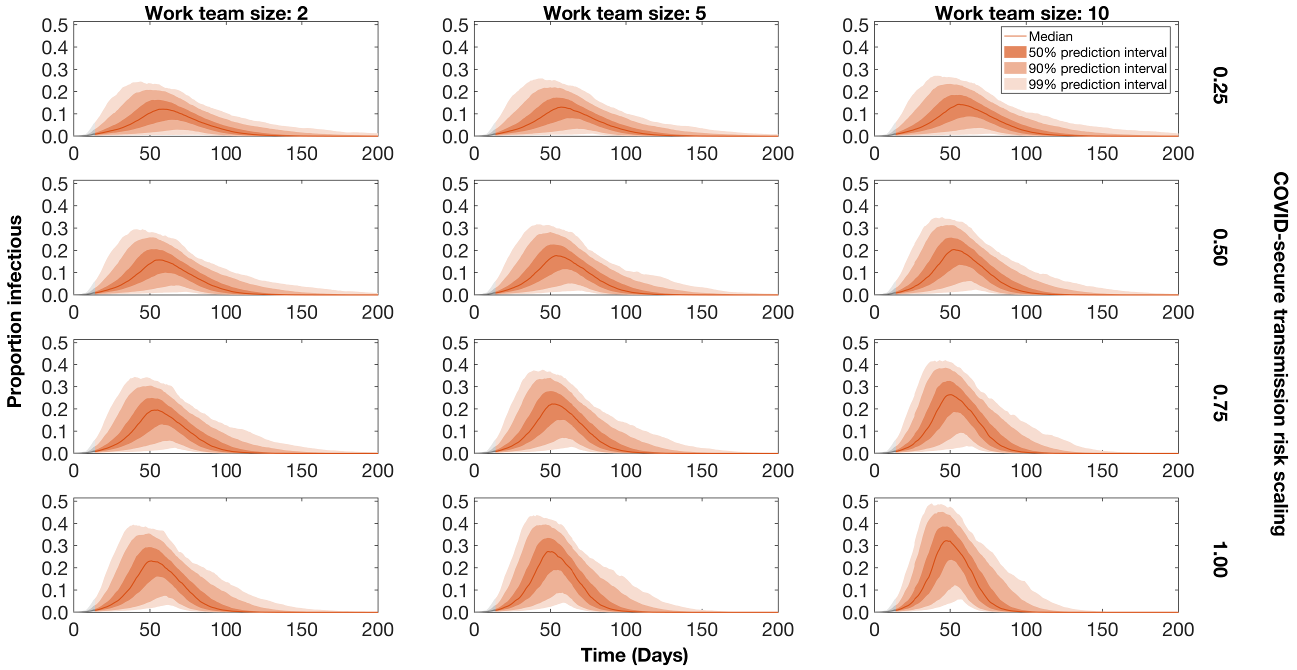

**Fig. S16: Prevalence temporal profiles and the under differing COVID-secure workplace assumptions in the absence of isolation, test and trace.** We display outputs for combinations of, from day 15, work team sizes being capped at two, five or 10 people, paired with scaling the transmission risk in COVID-secure workplaces by 0.25, 0.50, 0.75 or 1.00, respectively. We assumed all individuals were at the workplace five days a week (Monday to Friday). Traces and regions in grey correspond to the period where no interventions were in place (up to day 15). The solid line gives the median trace. Filled regions depict the 50%, 90% and 99% prediction intervals (with dark, moderate and light shading, respectively).

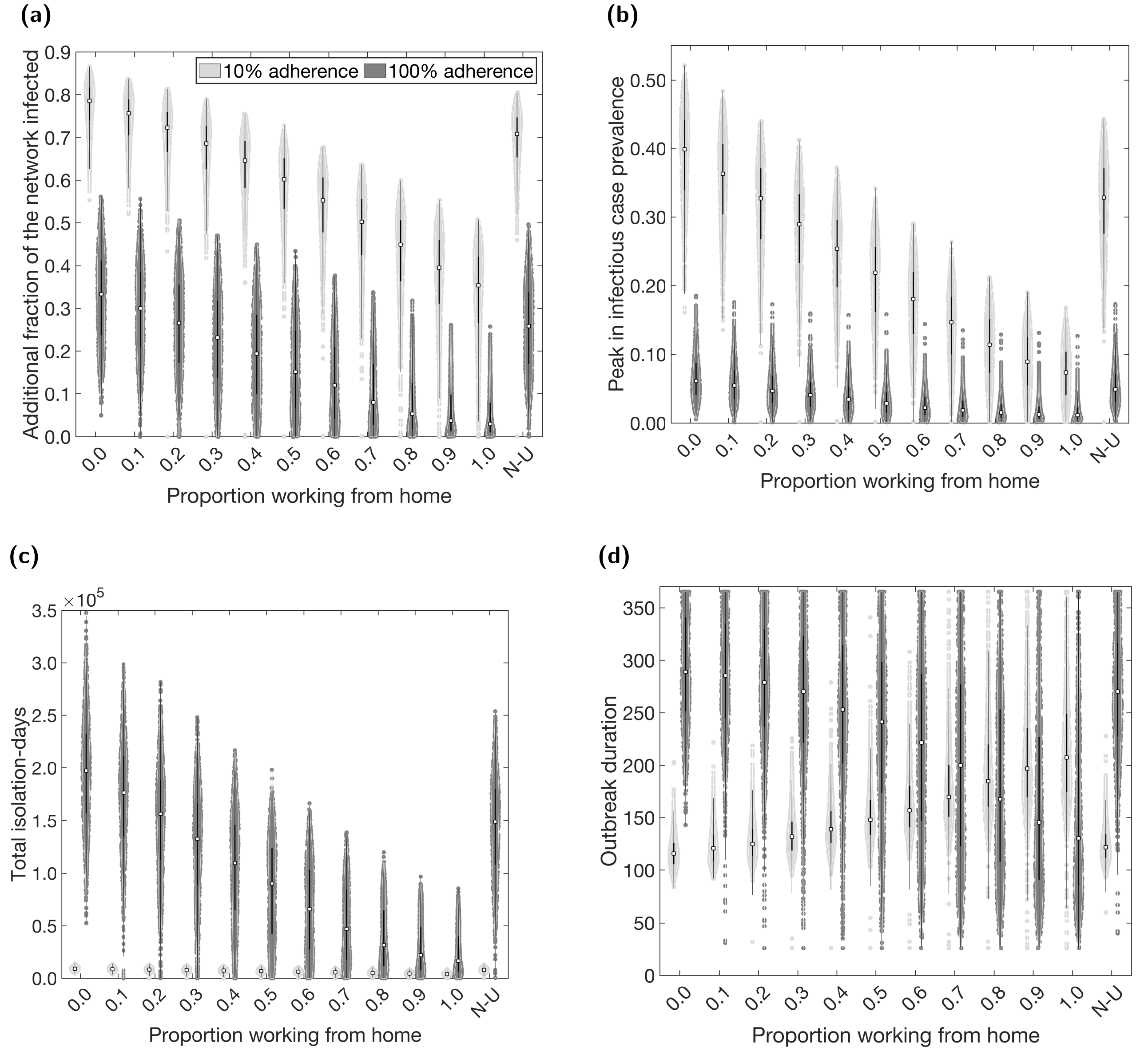

**Fig. S17: Case and isolation summary statistics under differing fractions of workers working from home and levels of adherence to testing, contact tracing and isolation.** We introduced NPIs from day 15 onwards, with varying proportions of the workforce working from home. N-U corresponds to non-uniform proportions working from home across the work sectors (see Table 5). We compare two scenarios of adherence to isolation and test-and-trace measures, 10% (light shaded violins) and 100% (dark shaded violins). Outputs are summarised from 1,000 simulations (20 runs per network for 50 separate network realisations). The white markers denote medians and solid black lines span the 25th to 75th percentiles. We give central and 95% prediction intervals in Table S9. **(a)** Additional proportion of the population that were infectious post introduction of NPIs (day 15 onwards). **(b)** Peak in infectious case prevalence. **(c)** Total isolation-days. **(d)** Outbreak duration.

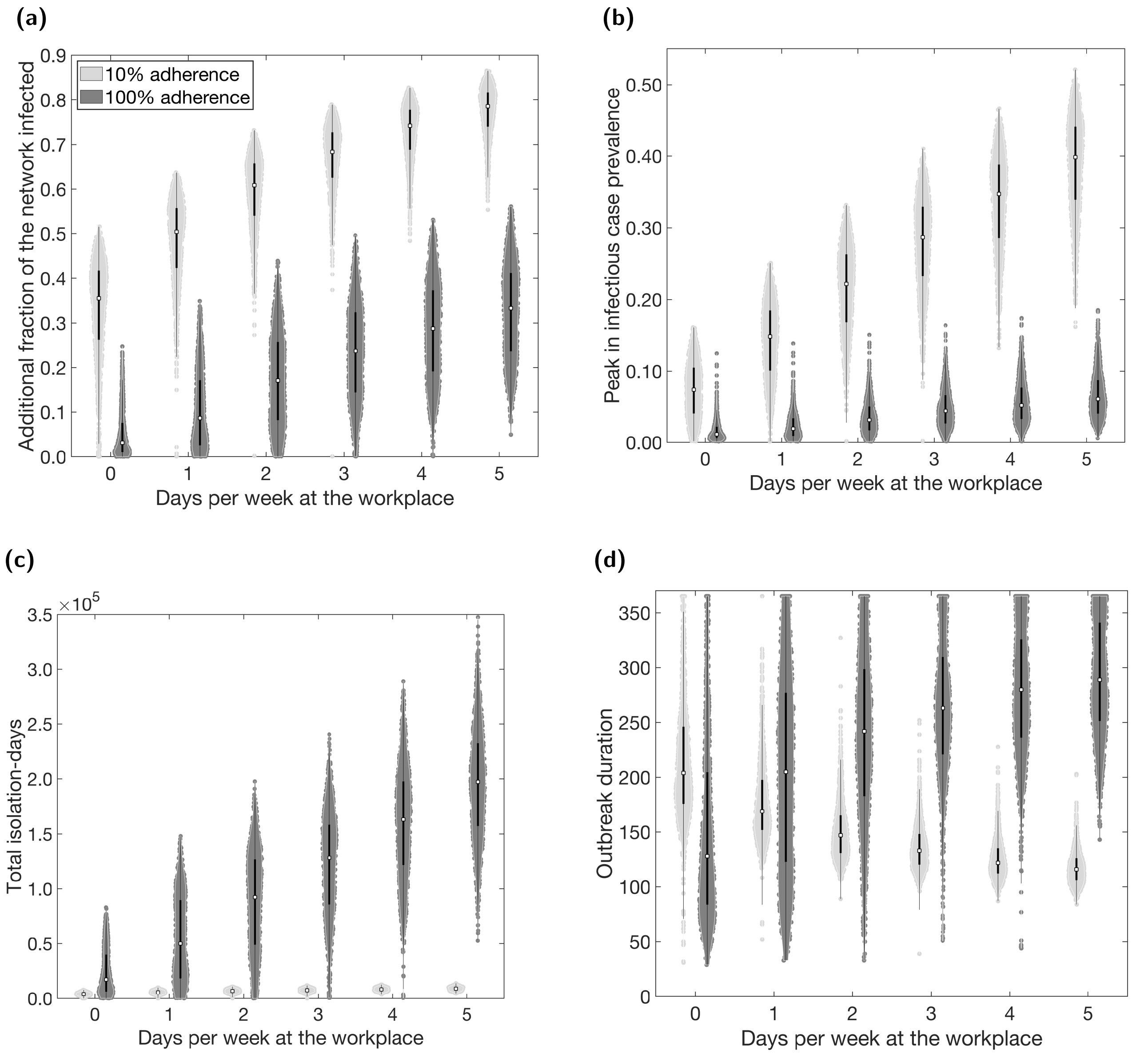

**Fig. S18: Case and isolation summary statistics under synchronous worker patterns and levels of adherence to testing, contact tracing and isolation.** We introduced NPIs from day 15 onwards and tested synchronous worker patterns for a range of days spent at the workplace. We compare two scenarios of adherence to isolation and test-and-trace measures, 10% (light shaded violins) and 100% (dark shaded violins). In all panels, we summarise outputs from 1,000 simulations (with 20 runs per network, for 50 network realisations). We assumed an adherence of 70% in all runs. The white markers denote medians and solid black lines span the 25th to 75th percentiles. We give central and 95% prediction intervals in Table S10. **(a)** Additional proportion of the population that were infectious post introduction of NPIs (day 15 onwards). **(b)** Peak in infectious case prevalence. **(c)** Total isolation-days. **(d)** Outbreak duration.

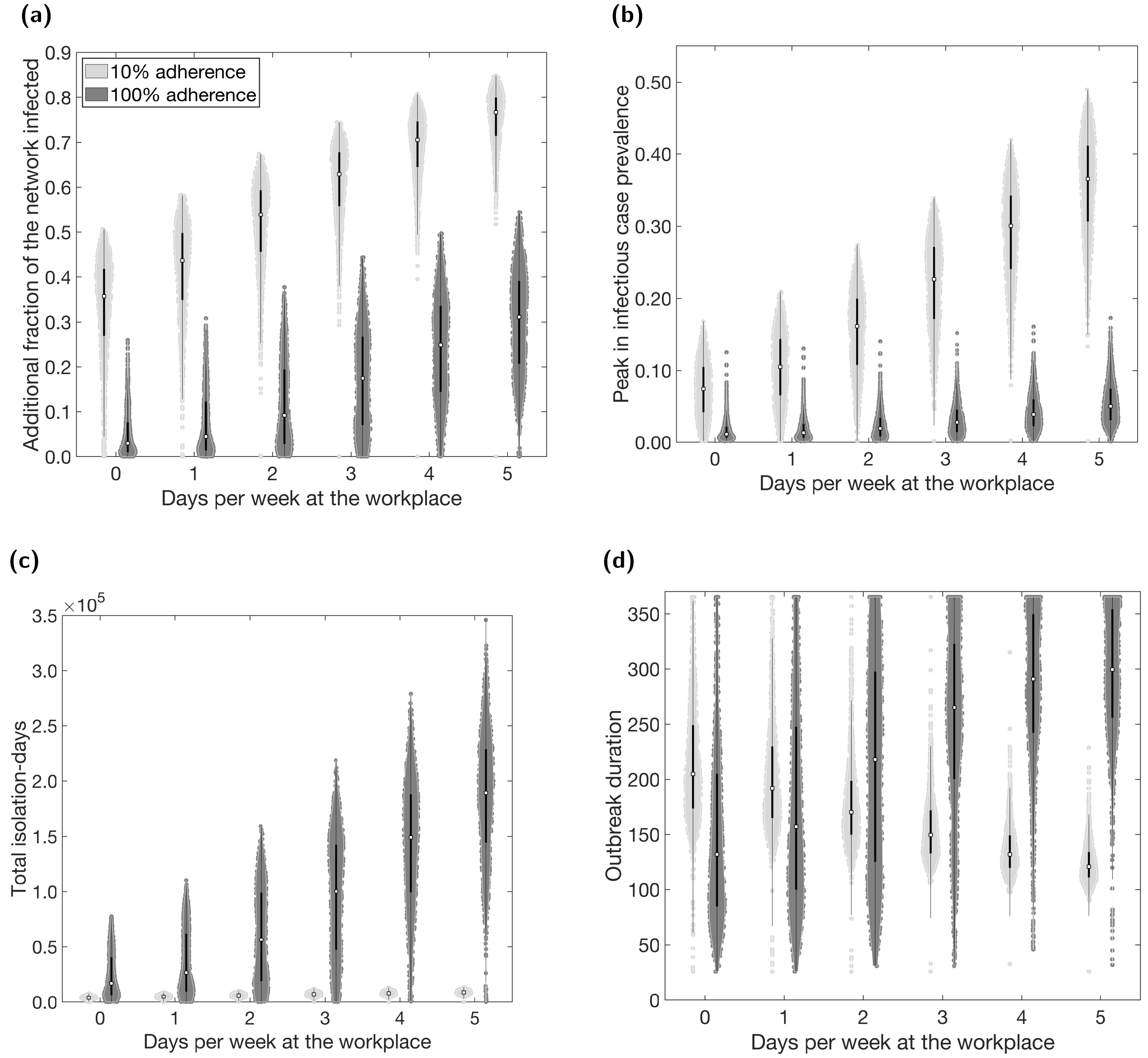

**Fig. S19: Case and isolation summary statistics under asynchronous worker patterns and levels of adherence to testing, contact tracing and isolation.** We introduced NPIs from day 15 onwards and tested asynchronous worker patterns for a range of days spent at the workplace. We compare two scenarios of adherence to isolation and test-and-trace measures, 10% (light shaded violins) and 100% (dark shaded violins). In all panels, we summarise outputs from 1,000 simulations (with 20 runs per network, for 50 network realisations). We assumed an adherence of 70% in all runs. The white markers denote medians and solid black lines span the 25th to 75th percentiles. We give central and 95% prediction intervals in Table S10. **(a)** Additional proportion of the population that were infectious post introduction of NPIs (day 15 onwards). **(b)** Peak in infectious case prevalence. **(c)** Total isolation-days. **(d)** Outbreak duration.

### 6 Additional tables

**Table S4:** Summary statistics for case, isolation and outbreak duration measures under differing strengths of interventions. We present median estimates and give 95% prediction intervals in parentheses, produced from 1,000 simulation replicates. We express the results for proportion infectious (day 15 onwards) and peak in infectious case prevalence to 2 d.p., total isolation-days to the nearest hundred and outbreak duration to the nearest integer.

| | | Probability | | | | | | | | | | | |
|---|---|---|---|---|---|---|---|---|---|---|---|---|---|
| **Variable** | **Statistic** | **0.0** | **0.1** | **0.2** | **0.3** | **0.4** | **0.5** | **0.6** | **0.7** | **0.8** | **0.9** | **1.0** | **N-U** |
| **Working from home** | **Proportion infectious (day 15 onwards)** | 0.51 (0.35,0.64) | 0.48 (0.32,0.61) | 0.45 (0.27,0.58) | 0.41 (0.24,0.55) | 0.37 (0.19,0.51) | 0.33 (0.14,0.48) | 0.29 (0.08,0.44) | 0.24 (0.03,0.41) | 0.20 (0.01,0.36) | 0.16 (0.01,0.32) | 0.13 (0.01,0.29) | 0.43 (0.28,0.56) |
| | **Peak in infectious case prevalence** | 0.16 (0.08,0.25) | 0.14 (0.07,0.23) | 0.12 (0.05,0.21) | 0.11 (0.04,0.19) | 0.09 (0.03,0.17) | 0.07 (0.02,0.14) | 0.06 (0.01,0.12) | 0.05 (0.01,0.11) | 0.04 (0.01,0.09) | 0.03 (0.00,0.08) | 0.02 (0.00,0.08) | 0.13 (0.06,0.21) |
| | **Total isolation-days** | 127400 (86000,162700) | 118200 (76300,155600) | 109700 (66600,146600) | 102100 (58500,137900) | 92300 (44800,130700) | 83100 (34200,119900) | 72000 (20800,109700) | 61500 (7300,98500) | 51400 (4300,88600) | 41300 (2600,78000) | 33500 (2000,69600) | 104100 (64300,139000) |
| | **Outbreak duration (days)** | 186 (140,272) | 191 (144,281) | 196 (145,290) | 200 (149,309) | 206 (154,325) | 214 (157,343) | 220 (152,365) | 230 (128,365) | 224 (113,365) | 218 (98,359) | 211 (81,365) | 190 (144,281) |
| **Adherence** | **Proportion infectious (day 15 onwards)** | 0.80 (0.66,0.87) | 0.78 (0.64,0.85) | 0.75 (0.61,0.82) | 0.72 (0.56,0.79) | 0.67 (0.52,0.75) | 0.62 (0.47,0.72) | 0.57 (0.42,0.68) | 0.51 (0.35,0.64) | 0.45 (0.29,0.59) | 0.39 (0.22,0.56) | 0.33 (0.13,0.51) | N/A |
| | **Peak in infectious case prevalence** | 0.42 (0.26,0.52) | 0.40 (0.24,0.49) | 0.37 (0.22,0.46) | 0.33 (0.19,0.42) | 0.28 (0.16,0.37) | 0.24 (0.14,0.33) | 0.20 (0.11,0.29) | 0.16 (0.08,0.25) | 0.12 (0.06,0.21) | 0.09 (0.04,0.18) | 0.07 (0.02,0.15) | N/A |
| | **Total isolation-days** | 0.00 (0.00, 0.00) | 8900 (5400,12000) | 22500 (14500,28900) | 39000 (26500,49800) | 58300 (40300,73200) | 79300 (54900,98800) | 102100 (70600,128900) | 127400 (86000,162700) | 152900 (100200,203400) | 176300 (107800,248100) | 193000 (98100,291900) | N/A |
| | **Outbreak duration (days)** | 110 (90,150) | 111 (92,150) | 117 (97,159) | 125 (101,166) | 135 (108,182) | 146 (115,200) | 163 (127,232) | 186 (140,272) | 216 (159,321) | 249 (179,365) | 283 (182,365) | N/A |

**Table S5:** Summary statistics for case, isolation and outbreak duration measures dependent upon the type of workplace attendance schedule. We present median estimates and give 95% prediction intervals in parentheses, produced from 1,000 simulation replicates. We express the results for proportion infectious (day 15 onwards) and peak in infectious case prevalence to 2 d.p., total isolation-days to the nearest hundred and outbreak duration to the nearest integer.

| | | Days at workplace | | | | | |
|---|---|---|---|---|---|---|---|
| **Worker pattern** | **Statistic** | **0** | **1** | **2** | **3** | **4** | **5** |
| **Synchronised** | **Proportion infectious (day 15 onwards)** | 0.13 (0.01,0.29) | 0.25 (0.03,0.40) | 0.34 (0.16,0.49) | 0.42 (0.25,0.55) | 0.46 (0.31,0.60) | 0.51 (0.35,0.64) |
| | **Peak in infectious case prevalence** | 0.02 (0.00,0.08) | 0.05 (0.01,0.11) | 0.08 (0.02,0.15) | 0.11 (0.04,0.19) | 0.13 (0.06,0.22) | 0.16 (0.08,0.25) |
| | **Total isolation-days** | 33500 (1800,68300) | 62700 (10000,101700) | 82400 (36700,118800) | 97700 (54300,132700) | 113900 (71700,149100) | 127400 (86000,162700) |
| | **Outbreak duration (days)** | 208 (74,365) | 228 (146,365) | 214 (155,329) | 201 (150,310) | 191 (145,283) | 186 (140,272) |
| **Asynchronised** | **Proportion infectious (day 15 onwards)** | 0.13 (0.01,0.29) | 0.19 (0.01,0.35) | 0.27 (0.04,0.43) | 0.35 (0.15,0.51) | 0.42 (0.25,0.57) | 0.48 (0.32,0.62) |
| | **Peak in infectious case prevalence** | 0.02 (0.00,0.08) | 0.03 (0.00,0.09) | 0.05 (0.01,0.11) | 0.07 (0.02,0.15) | 0.11 (0.04,0.19) | 0.14 (0.06,0.22) |
| | **Total isolation-days** | 34100 (2000,68700) | 48700 (3100,85600) | 69600 (7900,111600) | 91800 (38500,131100) | 111700 (64000,150700) | 126400 (82600,162400) |
| | **Outbreak duration (days)** | 216 (77,365) | 228 (100,365) | 238 (145,365) | 222 (164,358) | 210 (153,321) | 194 (147,298) |

**Table S6:** Summary statistics for case, isolation and outbreak duration measures dependent upon COVID-secure work interventions. We present median estimates and give 95% prediction intervals in parentheses, produced from 1,000 simulation replicates. We express the results for proportion infectious (day 15 onwards) and peak in infectious case prevalence to 2 d.p., total isolation-days to the nearest hundred and outbreak duration to the nearest integer.

| | | Work contact transmission scaling | | | |
|---|---|---|---|---|---|
| **Statistics** | **Work team size** | **0.25** | **0.50** | **0.75** | **1.00** |
| **Proportion infectious (day 15 onwards)** | **2** | 0.21 (0.02,0.37) | 0.28 (0.04,0.44) | 0.34 (0.09,0.50) | 0.40 (0.17,0.55) |
| | **5** | 0.23 (0.02,0.38) | 0.31 (0.07,0.46) | 0.38 (0.14,0.54) | 0.45 (0.24,0.60) |
| | **10** | 0.24 (0.03,0.40) | 0.34 (0.11,0.49) | 0.43 (0.22,0.57) | 0.50 (0.32,0.64) |
| **Peak in infectious case prevalence** | **2** | 0.04 (0.01,0.10) | 0.05 (0.01,0.12) | 0.06 (0.01,0.14) | 0.08 (0.02,0.17) |
| | **5** | 0.04 (0.01,0.10) | 0.06 (0.01,0.12) | 0.07 (0.02,0.16) | 0.10 (0.03,0.19) |
| | **10** | 0.04 (0.01,0.10) | 0.06 (0.01,0.14) | 0.09 (0.03,0.17) | 0.12 (0.05,0.22) |
| **Total isolation-days** | **2** | 69400 (6900,114800) | 82200 (14600,127300) | 93400 (25900,139200) | 103600 (44400,145100) |
| | **5** | 76700 (6500,124600) | 93500 (20800,141300) | 107600 (42700,153400) | 118700 (61200,160300) |
| | **10** | 87500 (9900,140600) | 109200 (36000,160200) | 124400 (62400,170500) | 134800 (83700,176600) |
| **Outbreak duration (days)** | **2** | 235 (124,365) | 241 (156,365) | 236 (164,365) | 235 (165,365) |
| | **5** | 241 (122,365) | 246(163,365) | 236 (164,365) | 222 (159,348) |
| | **10** | 244 (131,365) | 247 (170,365) | 229 (163,356) | 212 (150,333) |

**Table S7:** Epidemiological summary statistics when COVID-secure workplace interventions are active with a fixed team size of 5, varying transmission risk and for three different scenarios of adherence to test, trace and isolate measures (0%, 70% and 100% of the population). We give estimates of relative attack rate (from day 15), relative peak infectious proportion and relative outbreak duration for simulations including COVID-secure workplace measures compared to simulations without COVID-secure workplace measures (but with equivalent parameters and other interventions). We also provide the absolute values for each metric. We state median estimates and give 95% prediction intervals in parentheses, produced from 1,000 simulation replicates. We express all results to 2 d.p.

| | | | **Work contact transmission scaling** | | | |
|---|---|---|---|---|---|---|
| **Statistics** | **Adherence to test, trace, isolate** | **Reference value (median with no COVID-secure workplace intervention)** | **0.25** | **0.50** | **0.75** | **1.00** |
| **Relative attack rate (from day 15)** | **0%** | 0.80 | 0.65 (0.34,0.79) | 0.76 (0.47,0.89) | 0.86 (0.59,0.98) | 0.94 (0.70,1.04) |
| | **70%** | 0.51 | 0.44 (0.03,0.75) | 0.61 (0.13,0.91) | 0.75 (0.28,1.06) | 0.88 (0.47,1.18) |
| | **100%** | 0.33 | 0.19 (0.01,0.82) | 0.36 (0.01,1.08) | 0.57 (0.01,1.30) | 0.79 (0.02,1.46) |
| **Relative peak infectious proportion** | **0%** | 0.42 | 0.36 (0.09,0.58) | 0.49 (0.17,0.73) | 0.62 (0.25,0.87) | 0.75 (0.35,1.00) |
| | **70%** | 0.16 | 0.25 (0.03,0.62) | 0.35 (0.06,0.79) | 0.48 (0.11,1.00) | 0.64 (0.18,1.22) |
| | **100%** | 0.07 | 0.23 (0.04,0.97) | 0.30 (0.04,1.11) | 0.42 (0.04,1.38) | 0.55 (0.06,1.64) |
| **Relative outbreak duration** | **0%** | 109 days | 1.51 (1.10,2.61) | 1.40 (1.06,2.22) | 1.28 (0.99,1.93) | 1.19 (0.93,1.79) |
| | **70%** | 186 days | 1.30 (0.66,1.96) | 1.32 (0.88,1.96) | 1.27 (0.88,1.96) | 1.19 (0.85,1.87) |
| | **100%** | 283 days | 0.67 (0.19,1.29) | 0.88 (0.21,1.29) | 0.99 (0.27,1.29) | 1.05 (0.31,1.29) |
| **Proportion infectious (day 15 onwards)** | **0%** | — | 0.52 (0.27,0.63) | 0.61 (0.37,0.72) | 0.69 (0.48,0.79) | 0.76 (0.56,0.84) |
| | **70%** | — | 0.23 (0.02,0.38) | 0.31 (0.07,0.46) | 0.38 (0.14,0.54) | 0.45 (0.24,0.60) |
| | **100%** | — | 0.06 (0.00,0.27) | 0.12 (0.00,0.36) | 0.19 (0.00,0.43) | 0.26 (0.01,0.48) |
| **Peak in infectious case prevalence** | **0%** | — | 0.15 (0.04,0.25) | 0.21 (0.07,0.31) | 0.26 (0.10,0.37) | 0.32 (0.15,0.43) |
| | **70%** | — | 0.04 (0.01,0.10) | 0.06 (0.01,0.12) | 0.07 (0.02,0.16) | 0.10 (0.03,0.19) |
| | **100%** | — | 0.02 (0.00,0.07) | 0.02 (0.00,0.08) | 0.03 (0.00,0.09) | 0.04 (0.00,0.11) |
| **Outbreak duration (days)** | **0%** | — | 165 (120,284) | 153 (116,242) | 140 (108,211) | 130 (101,196) |
| | **70%** | — | 241 (122,365) | 246 (163,365) | 236 (164,365) | 222 (159,348) |
| | **100%** | — | 189 (54,365) | 248 (60,365) | 280 (77,365) | 297 (89,365) |

**Table S8:** Epidemiological summary statistics when COVID-secure workplace interventions were active with a fixed scaling of transmission risk of 0.5, varying work team size and for three different scenarios of adherence to test, trace and isolate measures (0%, 70% and 100% of the population). We give estimates of relative attack rate (from day 15), relative peak infectious proportion and relative outbreak duration for simulations including COVID-secure workplace measures compared to simulations without COVID-secure workplace measures (but with equivalent parameters and other interventions). We also provide the absolute values for each metric. We state median estimates and give 95% prediction intervals in parentheses, produced from 1,000 simulation replicates. We express all results to 2 d.p.

| | | | **Work team size** | | |
|---|---|---|---|---|---|
| **Statistics** | **Adherence to test, trace, isolate** | **Reference value (median with no COVID-secure workplace intervention)** | **2** | **5** | **10** |
| **Relative attack rate (from day 15)** | **0%** | 0.80 | 0.72 (0.41,0.85) | 0.76 (0.47,0.89) | 0.82 (0.53,0.94) |
| | **70%** | 0.51 | 0.55 (0.09,0.86) | 0.61 (0.13,0.91) | 0.67 (0.21,0.97) |
| | **100%** | 0.33 | 0.30 (0.01,0.98) | 0.36 (0.01,1.08) | 0.45 (0.01,1.16) |
| **Relative peak infectious proportion** | **0%** | 0.42 | 0.44 (0.14,0.66) | 0.49 (0.17,0.73) | 0.56 (0.21,0.80) |
| | **70%** | 0.16 | 0.32 (0.05,0.74) | 0.35 (0.06,0.79) | 0.41 (0.08,0.87) |
| | **100%** | 0.07 | 0.28 (0.04,1.04) | 0.30 (0.04,1.11) | 0.35 (0.04,1.19) |
| **Relative outbreak duration** | **0%** | 109 days | 1.44 (1.08,2.35) | 1.40 (1.06,2.22) | 1.33 (1.00,2.11) |
| | **70%** | 186 days | 1.30 (0.84,1.96) | 1.32 (0.88,1.96) | 1.33 (0.91,1.96) |
| | **100%** | 283 days | 0.80 (0.22,1.29) | 0.88 (0.21,1.29) | 0.95 (0.24,1.29) |
| **Proportion infectious (day 15 onwards)** | **0%** | — | 0.58 (0.33,0.69) | 0.61 (0.37,0.72) | 0.66 (0.43,0.76) |
| | **70%** | — | 0.28 (0.04,0.44) | 0.31 (0.07,0.46) | 0.34 (0.11,0.49) |
| | **100%** | — | 0.10 (0.00,0.33) | 0.12 (0.00,0.36) | 0.15 (0.00,0.38) |
| **Peak in infectious case prevalence** | **0%** | — | 0.19 (0.06,0.28) | 0.21 (0.07,0.31) | 0.24 (0.09,0.34) |
| | **70%** | — | 0.05 (0.01,0.12) | 0.06 (0.01,0.12) | 0.06 (0.01,0.14) |
| | **100%** | — | 0.02 (0.00,0.07) | 0.02 (0.00,0.08) | 0.02 (0.00,0.08) |
| **Outbreak duration (days)** | **0%** | — | 157 (118,256) | 153 (116,242) | 145 (109,231) |
| | **70%** | — | 241 (156,365) | 246 (163,365) | 247 (170,365) |
| | **100%** | — | 226 (61,365) | 248 (60,365) | 268 (67,365) |

**Table S9:** Summary statistics for case, isolation and outbreak duration measures under differing probabilities of working from home and adherences to testing, engagement with contact tracing and isolation. We present median estimates and give 95% prediction intervals in parentheses, produced from 1,000 simulation replicates. We express the results for total isolation-days to the nearest hundred, outbreak duration to the nearest integer and all remaining values to 2 d.p.

| | | Probability | | | | | | | | | | | |
|---|---|---|---|---|---|---|---|---|---|---|---|---|---|
| **Adherence to test, trace, isolate** | **Statistic** | **0.0** | **0.1** | **0.2** | **0.3** | **0.4** | **0.5** | **0.6** | **0.7** | **0.8** | **0.9** | **1.0** | **N-U** |
| **10%** | **Proportion infectious (day 15 onwards)** | 0.79 (0.63,0.85) | 0.76 (0.60,0.83) | 0.72 (0.55,0.80) | 0.69 (0.50,0.77) | 0.65 (0.45,0.74) | 0.60 (0.38,0.70) | 0.55 (0.31,0.66) | 0.50 (0.25,0.62) | 0.45 (0.16,0.57) | 0.40 (0.03,0.52) | 0.35 (0.03,0.48) | 0.71 (0.54,0.79) |
| | **Peak in infectious case prevalence** | 0.40 (0.24,0.49) | 0.36 (0.21,0.46) | 0.33 (0.18,0.42) | 0.29 (0.15,0.39) | 0.25 (0.11,0.35) | 0.22 (0.09,0.31) | 0.18 (0.06,0.27) | 0.15 (0.04,0.23) | 0.11 (0.02,0.19) | 0.09 (0.01,0.17) | 0.07 (0.00,0.14) | 0.33 (0.18,0.42) |
| | **Total isolation-days** | 9100 (5600,12500) | 8700 (5100,12100) | 8300 (4700,11600) | 7800 (4300,11400) | 7400 (3900,10700) | 6800 (3300,10200) | 6300 (2800,9700) | 5700 (2200,9200) | 5000 (1400,8400) | 4500 (400,7700) | 4100 (300,7200) | 8000 (4600,11300) |
| | **Outbreak duration (days)** | 116 (94,158) | 121 (98,166) | 125 (101,176) | 132 (104,189) | 139 (108,198) | 148 (115,226) | 157 (121,250) | 170 (124,293) | 185 (131,330) | 197 (130,363) | 208 (136,365) | 122 (99,170) |
| **100%** | **Proportion infectious (day 15 onwards)** | 0.33 (0.14,0.52) | 0.30 (0.09,0.50) | 0.27 (0.06,0.47) | 0.23 (0.01,0.43) | 0.19 (0.01,0.40) | 0.15 (0.00,0.37) | 0.12 (0.00,0.33) | 0.08 (0.00,0.29) | 0.05 (0.00,0.25) | 0.04 (0.00,0.22) | 0.03 (0.00,0.19) | 0.26 (0.07,0.45) |
| | **Peak in infectious case prevalence** | 0.06 (0.02,0.14) | 0.05 (0.01,0.13) | 0.05 (0.01,0.12) | 0.04 (0.00,0.10) | 0.03 (0.00,0.10) | 0.03 (0.00,0.08) | 0.02 (0.00,0.08) | 0.02 (0.00,0.07) | 0.02 (0.00,0.06) | 0.01 (0.00,0.06) | 0.01 (0.00,0.06) | 0.05 (0.01,0.12) |
| | **Total isolation-days** | 197400 (100900,295900) | 176700 (68300,270800) | 156800 (47300,249800) | 132800 (11700,214800) | 109900 (5500,190000) | 90000 (3900,164200) | 65900 (2200,139900) | 47200 (1500,119600) | 31500 (1200,97900) | 22000 (1100,81500) | 16600 (900,69300) | 149000 (50800,229300) |
| | **Outbreak duration (days)** | 289 (193,365) | 286 (168,365) | 279 (150,365) | 270 (107,365) | 253 (82,365) | 242 (65,365) | 222 (64,365) | 200 (55,365) | 168 (55,365) | 146 (49,365) | 131 (47,365) | 270 (151,365) |

**Table S10:** Summary statistics for case, isolation and outbreak duration measures under differing worker patterns and adherences to testing, engagement with contact tracing and isolation. We present median estimates and give 95% prediction intervals in parentheses, produced from 1,000 simulation replicates. We express the results for total isolation-days to the nearest hundred, outbreak duration to the nearest integer and all remaining values to 2 d.p.

| | | | Days at workplace | | | | | |
|---|---|---|---|---|---|---|---|---|
| **Worker pattern** | **Adherence to test, trace, isolate** | **Statistic** | **0** | **1** | **2** | **3** | **4** | **5** |
| **Synchronised** | **10%** | **Proportion infectious from day 15** | 0.36 (0.03,0.48) | 0.50 (0.26,0.61) | 0.61 (0.39,0.71) | 0.68 (0.50,0.77) | 0.74 (0.58,0.82) | 0.79 (0.63,0.85) |
| | | **Peak in infectious case prevalence** | 0.07 (0.01,0.14) | 0.15 (0.04,0.23) | 0.22 (0.09,0.31) | 0.29 (0.14,0.38) | 0.35 (0.19,0.44) | 0.40 (0.24,0.49) |
| | | **Total isolation-days** | 4100 (400,7200) | 5700 (2300,9100) | 6900 (3400,10200) | 7600 (4200,10900) | 8400 (5000,11900) | 9100 (5600,12500) |
| | | **Outbreak duration (days)** | 204 (131,365) | 169 (128,279) | 147 (115,219) | 133 (106,189) | 122 (100,172) | 116 (94,158) |
| | **100%** | **Proportion infectious from day 15** | 0.03 (0.00,0.19) | 0.09 (0.00,0.30) | 0.17 (0.01,0.38) | 0.24 (0.02,0.44) | 0.29 (0.08,0.49) | 0.33 (0.14,0.52) |
| | | **Peak in infectious case prevalence** | 0.01 (0.00,0.06) | 0.02 (0.00,0.07) | 0.03 (0.00,0.09) | 0.04 (0.01,0.11) | 0.05 (0.01,0.13) | 0.06 (0.02,0.14) |
| | | **Total isolation-days** | 17400 (900,70100) | 50200 (1700,123100) | 92400 (4800,166200) | 128700 (18000,207300) | 163700 (65100,254300) | 197400 (100900,295900) |
| | | **Outbreak duration (days)** | 128 (47,363) | 205 (56,365) | 242 (73,365) | 263 (115,365) | 280 (168,365) | 289 (193,365) |
| **Asynchronised** | **10%** | **Proportion infectious from day 15** | 0.36 (0.03,0.48) | 0.44 (0.15,0.56) | 0.54 (0.27,0.65) | 0.63 (0.41,0.73) | 0.71 (0.52,0.79) | 0.77 (0.61,0.84) |
| | | **Peak in infectious case prevalence** | 0.07 (0.01,0.15) | 0.11 (0.01,0.19) | 0.16 (0.04,0.25) | 0.23 (0.09,0.32) | 0.30 (0.15,0.39) | 0.37 (0.20,0.46) |
| | | **Total isolation-days** | 4100 (300,7200) | 5000 (1300,8200) | 6100 (2400,9500) | 7300 (3700,10700) | 8100 (4600,11600) | 8900 (5400,12300) |
| | | **Outbreak duration (days)** | 205 (134,365) | 192 (134,365) | 171 (123,304) | 150 (115,233) | 132 (106,195) | 121 (98,172) |
| | **100%** | **Proportion infectious from day 15** | 0.03 (0.00,0.18) | 0.05 (0.00,0.25) | 0.09 (0.00,0.32) | 0.17 (0.00,0.40) | 0.25 (0.02,0.46) | 0.31 (0.08,0.50) |
| | | **Peak in infectious case prevalence** | 0.01 (0.00,0.06) | 0.01 (0.00,0.06) | 0.02 (0.00,0.07) | 0.03 (0.00,0.09) | 0.04 (0.00,0.11) | 0.05 (0.01,0.13) |
| | | **Total isolation-days** | 17100 (900,68200) | 26800 (1200,94000) | 56600 (1800,137400) | 100300 (4400,187500) | 149300 (16400,245100) | 189500 (64000,291700) |
| | | **Outbreak duration (days)** | 132 (46,364) | 157 (50,365) | 218 (55,365) | 265 (69,365) | 291 (106,365) | 300 (153,365) |
